# Transient anti-interferon autoantibodies in the airways are associated with efficient recovery from COVID-19

**DOI:** 10.1101/2024.01.11.24301000

**Authors:** Benjamin R. Babcock, Astrid Kosters, Devon J. Eddins, Maria Sophia Baluyot Donaire, Sannidhi Sarvadhavabhatla, Vivian Pae, Fiona Beltran, Victoria W. Murray, Gurjot Gill, Guorui Xie, Brian S. Dobosh, Vincent D. Giacalone, Rabindra M. Tirouvanziam, Richard P. Ramonell, Scott A. Jenks, Ignacio Sanz, F. Eun-Hyung Lee, Nadia R. Roan, Sulggi A. Lee, Eliver E. B. Ghosn

**Affiliations:** Department of Medicine, Division of Immunology and Rheumatology, Lowance Center for Human Immunology, Emory University, Atlanta, GA; Department of Medicine, Division of HIV, Infectious Diseases & Global Medicine, University of California, San Francisco, CA; Department of Pediatrics, Emory University, Atlanta, GA; Department of Medicine, Division of Pulmonary, Allergy, Critical Care, and Sleep Medicine, Emory University, Atlanta, GA; Gladstone Institutes, San Francisco, CA; Department of Urology, University of California San Francisco, San Francisco, CA; Emory Vaccine Center, Emory National Primate Research Center, Emory University, Atlanta, GA

**Author notes:** **Corresponding Author Contact Info** Eliver E. B. Ghosn: Health Sciences Research Building-I, 1760 Haygood Dr. NE, E240, Atlanta, GA 30322, USA. The current affiliation for D.J.E is the National Center for Immunization and Respiratory Diseases (NCIRD), Centers for Disease Control and Prevention (CDC), Atlanta, GA. The current affiliation for R.P.R. is the Division of Pulmonary, Department of Medicine, Allergy and Critical Care Medicine, University of Pittsburgh School of Medicine, Pittsburgh, PA.

## Abstract

Pre-existing anti-interferon alpha (anti-IFN-α) autoantibodies in blood are associated with susceptibility to life-threatening COVID-19. However, it is unclear whether anti-IFN-α autoantibodies in the airways – the initial site of infection – can also determine disease outcomes. In this study, we developed a new multiparameter technology, flowBEAT, to quantify and profile the isotypes of anti-IFN-α and anti-SARS-CoV-2 antibodies in longitudinal samples collected over 20 months from the airway and matching blood of 129 donors with mild, moderate, and severe COVID-19. We found unexpectedly that nasal anti-IFN-α autoantibodies were induced post-infection onset in more than 70% of mild to moderate COVID-19 cases and associated with robust anti-SARS-CoV-2 immunity, fewer symptoms, and efficient recovery. Nasal anti-IFN-α autoantibodies followed the peak of host IFN-α production and waned with disease recovery, revealing a regulated balance between IFN-α and anti-IFN-α response. Notably, only a subset of mild to moderate patients progressed to develop systemic anti-IFN-α, which correlated with systemic inflammation and worsened symptoms. In contrast, patients with life-threatening COVID-19 sustained elevated anti-IFN-α in both airways and blood, coupled with uncontrolled viral load and IFN-α production. Our studies thereby reveal a novel protective role for nasal anti-IFN-α autoantibodies in the immunopathology of COVID-19 and, more broadly, suggest that anti-IFN-α may serve an important regulatory function to restore homeostasis following viral invasion of the respiratory mucosa.

## INTRODUCTION

Although most individuals are susceptible to respiratory SARS-CoV-2 infection, only a small group develops life-threatening, severe COVID-19 disease. We and others have shown that life-threatening COVID-19 is associated with an uncontrolled hyper-inflammatory response in the airways (1-4) rather than an uncontrolled viral load (1, 5). While these previous studies have revealed the hyper-inflammatory immune phenotypes (1-4) associated with life-threatening COVID-19, the protective mechanisms that restrict viral replication while preventing hyper-inflammation and cytokine release syndrome in the airways during natural recovery remain unclear.

Of the many inflammatory cytokines produced in the airways during SARS-CoV-2 infection, type-I interferons (IFNs) have been extensively studied due to their protective antiviral properties, particularly when produced early at the disease onset (6-9). However, type-I IFNs can also worsen symptoms during viral infections. Delayed or exacerbated IFN-α production in the airways of COVID-19 patients (8) or animal models of coronavirus infection (9-11) is pathologic and associated with increased disease severity. Thus, there is a contradiction wherein early IFN-α elicits antiviral protection while delayed or persistent IFN-α can trigger hyper-inflammation and worsening symptoms during viral infections.

Patients with pre-existing defects in IFN-α responses, including inborn genetic errors, are predisposed to life-threatening COVID-19 (12-14), supporting a protective role for IFN-α. Consistent with this, autoantibodies against IFN-α (anti-IFN-α) in the blood are associated with an increased risk of life-threatening disease (15-19). Paradoxically, increased and prolonged production of IFN-α in the airways is detrimental and associated with progression to severe COVID-19 (3, 8, 20-23), suggesting that a strictly regulated IFN-α response locally in the airway mucosa is instrumental in determining disease outcome. Indeed, controlled IFN-α production strictly limited to the early stages of infection appears beneficial and associated with efficient recovery from COVID-19 (8, 11). Therefore, an important unanswered question is whether a regulated balance between host IFN-α, anti-IFN-α autoantibodies, and anti-SARS-CoV-2 antibodies in the airway mucosa is necessary for efficient recovery during natural infection and whether dysregulation of this balance in the airways is detrimental, leading to life-threatening COVID-19.

In this study, we developed flowBEAT (flow cytometry-based Bead assay to detect Antigen-specific antibody isoTypes) to determine the longitudinal dynamics of anti-IFN-α autoantibodies and anti-SARS-CoV-2 in the airway sites of infection and matched blood, revealing their contribution to the progression of COVID-19 spanning from disease onset to full recovery. We show that anti-IFN-α autoantibodies in the airways are a common feature of efficient recovery from COVID-19, as they are transiently induced in patients with mild and moderate disease. Collectively, our studies reveal a coordinated and transient induction of anti-IFN-α and host IFN-α in the airways and establish that viral-induced anti-IFN-α in the nasal mucosa, but not in the blood is associated with fewer symptoms, robust anti-SARS-CoV-2 antibodies, and full recovery.

## RESULTS

### FlowBEAT reveals distinct anti-SARS-CoV-2 and anti-type I IFN autoantibody responses across tissues and disease state

To quantify the full breadth of antibody responses, including isotype usage and antigen specificity across the COVID-19 disease state, we developed a multiparameter assay called “flowBEAT” (flow cytometry-based Bead assay to detect Antigen-specific antibody isoTypes). FlowBEAT is a modular flow cytometry-based technology that measures 176 antibody parameters per sample, including eight human antibody isotypes (IgG1, IgG2, IgG3, IgG4, IgA1, IgA2, IgE, IgM) against a panel of 22 host and viral antigens (Supplemental Table 1), including host type-I interferons (anti-IFN-α2a and anti-IFN-ω) and SARS-CoV-2 structural (anti-spike, -nucleocapsid, -membrane, and - envelope) and non-structural (anti-NSPs and -ORFs) proteins. We applied FlowBEAT to paired airway and blood samples from COVID-19 patients (Fig. 1 and Supplemental Table 1).

**Figure 1.**
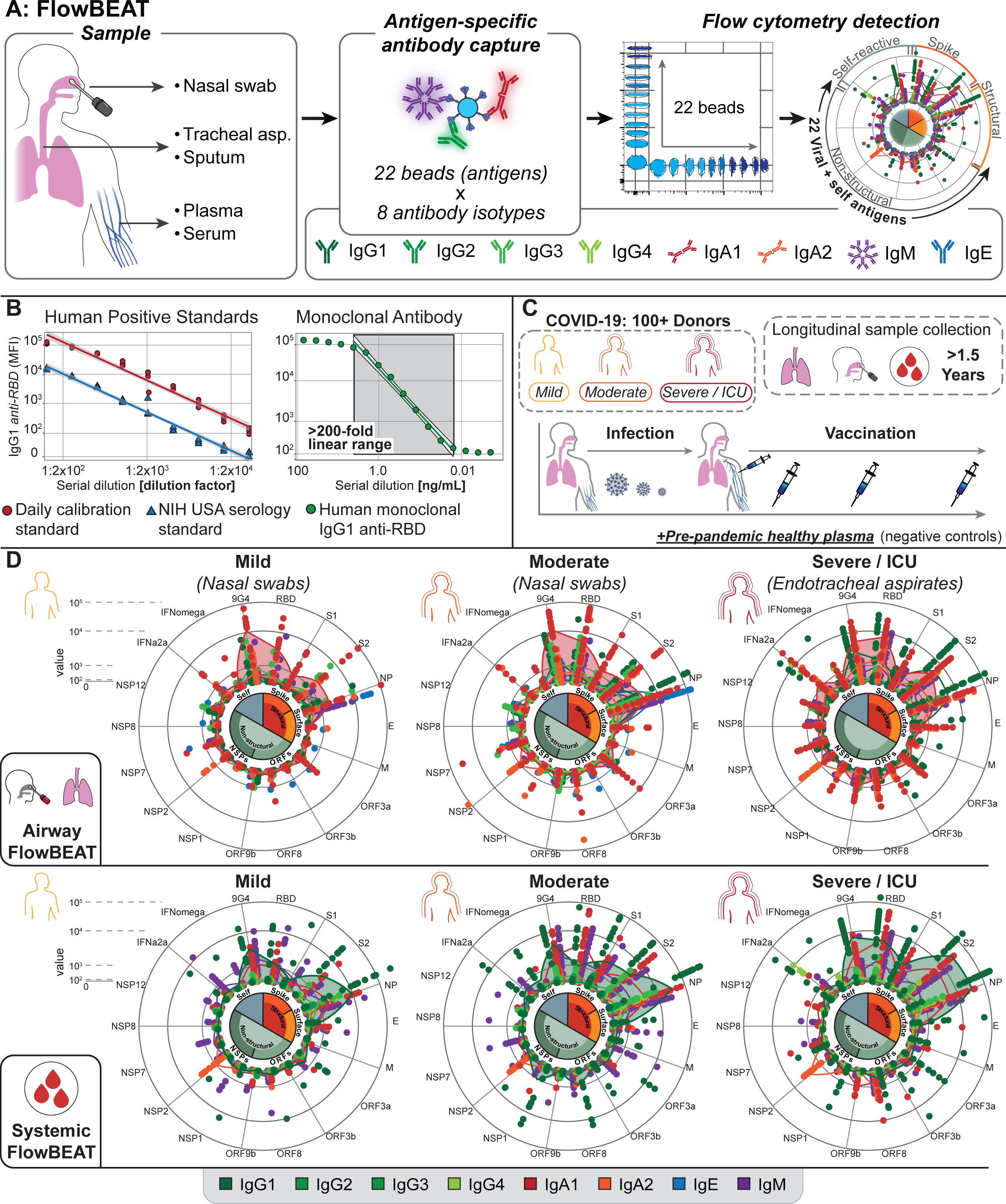
Multi-tissue validation of the flowBEAT assay and cohort summary. (**A**) Graphical overview of flowBEAT sampling from different tissues to reveal antibody isotype (color scheme 1) and antigen specificity (color scheme 2). (**B**) Dot plot showing the linear decrease of anti-RBD signal by flowBEAT serial dilution of seropositive NIH standards and monoclonal human IgG1 anti-RBD. (**C**) Graphical summary of study cohort spanning mild, moderate, and severe COVID-19 with multi-tissue collection and longitudinal follow-up. See cohort details in Tables S2-5. (**D**) Sunburst plots of flowBEAT showing the breadth of antibody response against SARS-CoV-2 and autoantigens in the airway and systemic samples, grouped by disease severity.

We first determined the specificity, sensitivity, and reproducibility of the flowBEAT antigen-coated beads. We established the linear range of the assay by generating replicate serial dilutions of mouse and human monoclonal antibodies, NIH COVID-19 human serology standard (24) (Fig. 1B), and pre-pandemic serum as negative controls (Fig. 1B and Supplemental Figs. 1-2). We determined the background signal of non-specific antibody binding using BSA-coated control beads, which were added to each tube during data collection and whose signals were used to establish the lower limits of detection of the assay (Methods).

**Figure 2.**
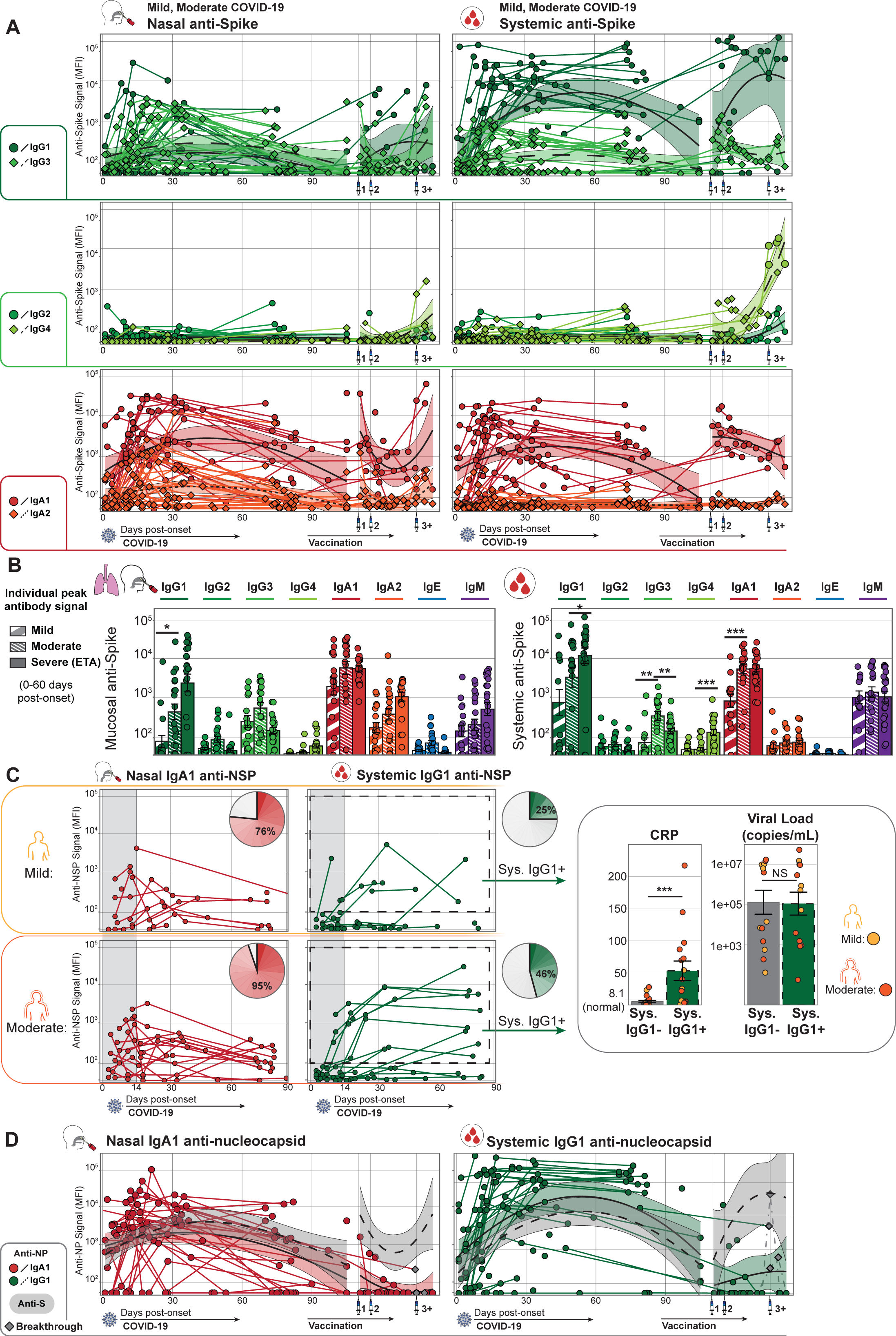
Longitudinal kinetics of anti-SARS-CoV-2 antibody isotype response associated with disease severity and vaccination in the airway mucosa and blood. (**A**) Longitudinal plots showing isotype-specific anti-Spike signal (Y-axis) throughout infection and up to four doses of mRNA vaccination (X-axis) in both the nasal mucosa and blood of mild and moderate COVID-19 patients. Solid lines connect longitudinal samples from individual patients, and a Loess regression showing isotype induction and wane is plotted on top with standard error as a shaded area. (**B**) Bar plots show the maximum anti-Spike signal by individual patients (points, one per patient) and are grouped by severity class (fill texture) and antibody isotype (color). Statistics by Wilcoxon rank-sum test. (**C**) Tissue-specific anti-NSP signature. 31/36 donors show nasal IgA1 anti-NSP (NSP1, NSP2, NSP12, ORF3a, ORF3b, and ORF8) within the first two weeks. Of these, 16/36 progress to develop systemic anti-NSP IgG1 after the first two weeks. Pie charts detail positive-detected donors, with shading corresponding to peak antibody level. Dashed lines represent gates to select donors who generate anti-NSP IgG1. Bar charts of CRP and peak viral load, with donors grouped by anti-NSP IgG1 status, points colored by severity group. Statistics by Wilcoxon rank-sum test. (**D**) Longitudinal plots showing induction and waning of anti-NP antibodies. Grey points highlight suspected viral exposures, identified by an increase in anti-NP signal. Grey Loess regression indicates anti-Spike signal as in (A).

Using flowBEAT, we measured the breadth of antibody responses in the airway mucosa and matching blood of 129 donors spanning from mild (n = 37), moderate (n = 33), and severe life-threatening (n = 23) COVID-19, sampled longitudinally by repeated collections throughout infection and subsequent vaccination. We also collected samples from infection-naïve donors (n = 36), including vaccinated and pre-pandemic donors, for a total of 505 unique samples (205 airway mucosal samples and 300 systemic serum/plasma samples, Fig. 1C and Supplemental Tables 2-4). FlowBEAT revealed that the amount and diversity of antibody response increased with disease severity, represented as peak antibody expression during acute infection (Fig 1D). As the disease severity increases from mild to severe COVID-19, new antibody isotypes and specificities were generated against SARS-CoV-2 non-structural proteins (NSP and ORF) and host IFNs (Fig. 1D). Thus, flowBEAT distinguished the breadth of antibody response against SARS-CoV-2 and host IFNs across tissues and disease state.

### Airway-specific antibody isotype and specificity against SARS-CoV-2 proteins distinguish mild, moderate, and severe disease

To better understand the breadth of antibody response associated with disease progression and recovery, we assessed the isotype and specificity of antibodies against SARS-CoV-2 proteins longitudinally and across disease states. We longitudinally sampled the nasal mucosa and blood starting at disease onset, determined as the date of polymerase-chain reaction (PCR)-confirmed infection, and through recovery and subsequent mRNA vaccination. In mild and moderate COVID-19 patients who recovered from the disease, IgG1, IgG3, and IgA1 were the predominant anti-spike (anti-S) antibody isotypes in both nasal mucosa and blood, reaching peak levels before 30 days post-onset (Fig. 2A). While most antibody isotypes were maintained for more than three months post-onset in blood, they quickly waned in the nasal mucosa after disease recovery, except for IgA1 which was still detectable in > 60% of mild/moderate-recovered donors three months post-onset (Fig. 2A). Interestingly, nasal IgA1 and IgE isotypes of anti-S and anti-nucleocapsid protein (anti-NP) pre-dated the detection of IgM by at least one week (Supplemental Fig. 3), likely representing pre-existing immunity to S and NP epitopes conserved between SARS-CoV-2 and common cold coronaviruses (25, 26).

We also found airway- and blood-specific antibody isotype and specificity associated with COVID-19 disease state. In the airway, the production of IgA2, often related to hyperinflammatory responses in the mucosa (27, 28), increased with disease severity and was highest within the endotracheal aspirates (ETA) of acutely-infected severe COVID-19 patients (Fig. 2B). In blood, while IgM was comparable across disease states, a sharp increase in the production of IgG1 and IgG4 anti-S associated with increased COVID-19 severity (Fig. 2B). High IgG3, which can suppress type-I IFN (29), exclusively distinguished patients with moderate to severe disease. While 17/28 of moderate cases of COVID-19 developed high systemic IgG3, only 4/24 of mild cases had detectable systemic IgG3 during acute infection (<100 days post-onset) (Fig. 2B).

Finally, we identified a nasal-specific IgA1 antibody signature against other SARS-CoV-2 structural and non-structural proteins (NSP and ORF), which emerged within two weeks of infection (Fig. 2C and Supplemental Fig. 4). Notably, only a subset of patients, primarily those with moderate but not mild disease, went on to develop an equivalent anti-NSP IgG1 signature in the blood after two weeks of infection. This progression from nasal IgA1 to systemic IgG1 anti-NSP was linked to an increased level of systemic inflammation at the peak of infection (Fig. 2C). Thus, the initial nasal-specific antiviral (anti-NSP/ORF) response is restricted to the airway site of infection and progresses to systemic response only in patients with increased disease severity.

In contrast to the relative stability of isotype usage, particularly IgA1 and IgG1, throughout SARS-CoV-2 infection and recovery, vaccination induced de novo IgG2 and IgG4 after two or more mRNA vaccine doses (Fig. 2A). Furthermore, the isotypes IgG3 and IgA2 anti-S, which we show were associated with disease severity (Fig. 2B), were not boosted by mRNA vaccination (Fig. 2A). Surprisingly, we found that nasal IgA1 and IgG1 anti-S (but not anti-NP) were boosted by distal intramuscular mRNA vaccination (Fig. 2D), suggesting a role for vaccination in supporting mucosal sterilizing immunity and preventing community transmission. Thus, flowBEAT revealed an anti-NSP/ORF signature that is restricted to the airway site of infection in mild-recovered individuals and identified IgA2, IgG3, and IgG4 as potential biomarkers of COVID-19 disease severity, while IgG2 was induced only by repeated vaccination.

### Viral-induced anti-IFN-α autoantibodies in the nasal mucosa are associated with lesser symptoms, robust anti-SARS-CoV-2 antibodies, and full recovery

To gain a better insight into the longitudinal dynamics between anti-SARS-CoV-2 antibodies and autoantibodies against type-I IFN associated with full recovery, we longitudinally assessed 36 of the mild and moderate COVID-19 patients for whom we collected matched nasal and blood samples throughout infection and recovery. We profiled multiple samples from disease onset through recovery and subsequent mRNA vaccination and linked the data to symptomology and other clinical features (Supplemental Table 3). We first investigated whether anti-type I IFN autoantibodies (anti-IFN-α2a and anti-IFN-ω) could be detected locally in the airways throughout the COVID-19 disease progression. To confirm the specificity of the anti-IFN autoantibodies in our flowBEAT assay and to compare them to other published studies (16, 19, 30, 31), we coated our assay beads with the same IFN-α2a and IFN-ω proteins previously shown to identify anti-IFN autoantibodies in COVID-19 patients (30). To distinguish background binding from antigen-specific signal, we performed serial dilutions of positive samples and spiked BSA-coated beads in positive samples to calculate non-specific antibody binding to our IFN-coated beads (Supplemental Fig. 2). We were able to identify anti-IFN responders and non-responders in all our COVID-19 cohorts.

Strikingly, we discovered a robust anti-IFN-α2a autoantibody response induced by SARS-CoV-2 soon after infection, which peaked within the first two weeks post-onset in the nasal mucosa (nasal anti-IFN-α2a detected in 72% or 26/36 of mild and moderately-infected patients longitudinally assessed from disease onset) (Fig. 3A). These new-onset nasal anti-IFN-α2a autoantibodies were transient and waned as patients recovered from symptomatic disease (Fig. 3A). Consistent with the anti-SARS-CoV-2 antibody isotype response described above, nasal anti-IFN-α2a was dominated by the IgA1 isotype in mild and moderate cases of COVID-19, while IgA2, often associated with mucosal hyperinflammation (27, 28), was detected at lower levels (Fig. 3A). Notably, the initial anti-IFN-α response was limited to the nasal mucosa and rarely progressed to a systemic response in the blood. Only 13 of the 36 donors with matched blood and nasal swabs developed blood anti-IFN-α, which appeared later (after two weeks post-onset) (Fig. 3C) and was dominated by IgG1 (Fig. 3C and Supplemental Fig. 4). However, when anti-IFN-α progressed to the blood, it persisted for over three months post-disease onset (Fig. 3C) and was more frequently detected in the blood of donors with moderate (10/24) as compared to mild (4/28) disease (Fig. 3D). We observed a similar pattern of anti-IFN-ω in the same donors, but at lower levels (Supplemental Fig. 4). Finally, in contrast to COVID-19 disease, mRNA vaccination did not induce anti-IFN-α autoantibodies (Figs. 3A, C), indicating that anti-IFN-α autoantibodies are a specific response to SARS-CoV-2 infection.

**Figure 3.**
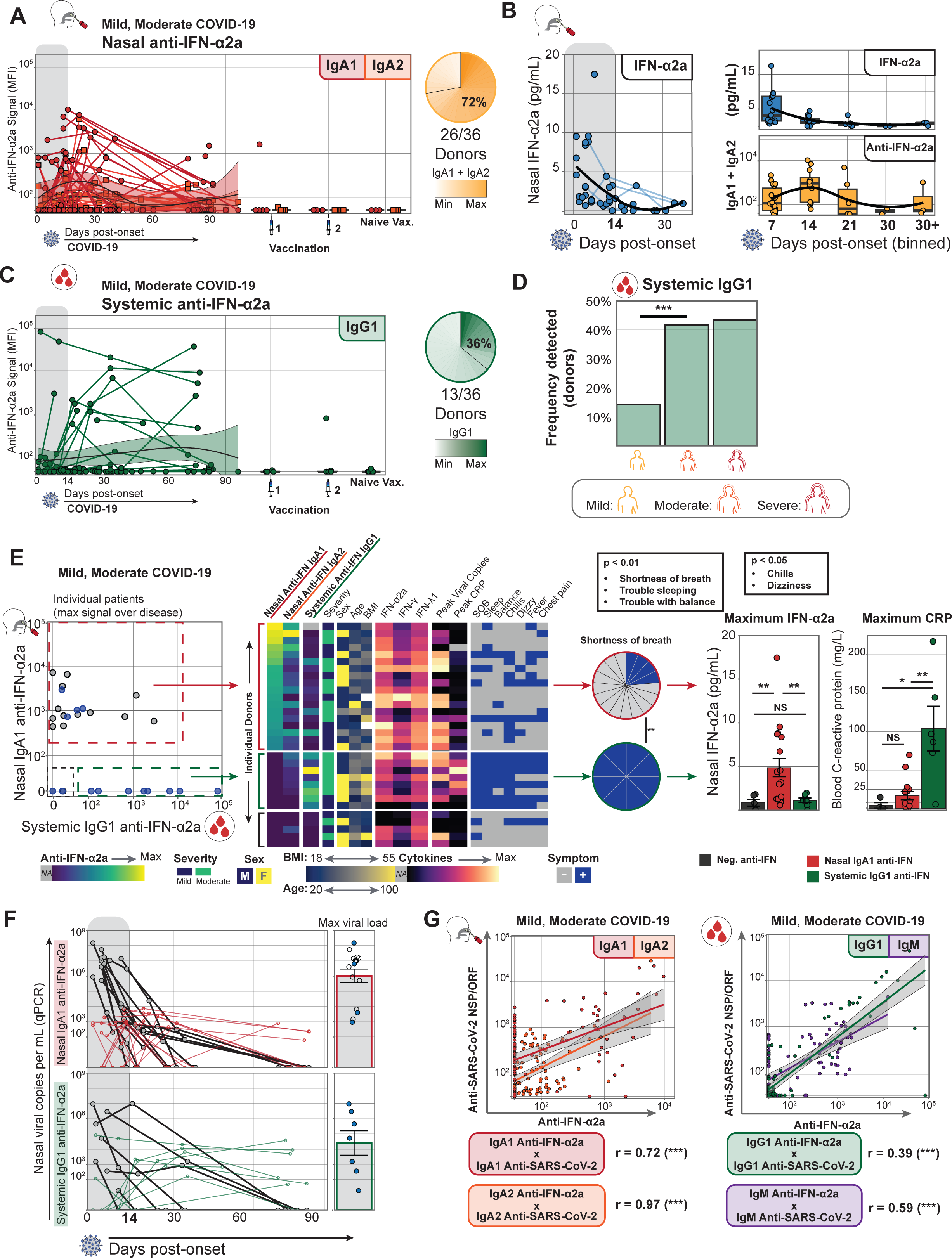
Transiently induced nasal anti-IFN-α autoantibodies are associated with less symptomology and efficient recovery from COVID-19. (**A**) Longitudinal plot showing the post-onset induction of nasal (IgA1, IgA2) anti-IFN-α2a autoantibodies in mild and moderate COVID-19. Days post-onset is shown on the X-axis with longitudinal samples from single donors linked by solid lines with population trends (Loess regression) plotted as solid black lines with shaded standard error. Pie charts show the frequency and maximum signal intensity of total mucosal IgA anti-IFN-α2a. (**B**) The longitudinal decline of nasal IFN-α2a cytokine plotted as pg/mL of nasal swab supernatant. Longitudinal plots binned by week, showing a decline of nasal IFN-α2a cytokine and an increase in nasal anti-IFN-α2a autoantibodies. (**C**) Longitudinal plot showing post-onset induction of systemic (IgG1) anti-IFN-α2a autoantibodies in mild and moderate COVID-19. Axes and Loess regression as in (A). Pie charts show the frequency and maximum signal intensity of total systemic IgG1 anti-IFN-α2a. (**D**) Frequency summary (percent detected) of systemic IgG1 anti-IFN-α2a grouped by COVID-19 severity (numbers of positive donors over total donors). Chi-square test, p < 0.001. (**E**) Dot plot of nasal IgA1 anti-IFN-α2a (Y-axis) and systemic anti-IFN-α2a IgG1 signals (X-axis). Points represent the patient’s maximum signal within the first six weeks post-onset. Patient populations are gated (dashed lines) as non-producers (black), nasal IgA1-producers (red), and systemic IgG1-producers (green). Heatmap of nasal IgA1 and IgA2, and blood IgG1 anti-IFN-α2a, patient demographic data, and features of local immune response and symptoms. Rows correspond to individual patients, while columns indicate features as labeled. Pie charts show the frequency of “shortness of breath” (SOB) symptoms between red and green gates, as well as the maximum signal for nasal IFN-α2a cytokine and systemic C-reactive protein (CRP) for each patient in the gated populations. Wilcoxon rank-sum test, p < 0.01 or p < 0.05, as indicated. (**F**) Longitudinal plots showing nasal viral load per mL (qPCR) and tissue-specific anti-IFN-α2a autoantibody kinetics. Groups are divided by red and green gates, as in (C). The bar chart summarizes the peak viral load. (**G**) Correlation plots between anti-IFN-α2a (X-axis) and composite anti-non-structural proteins (anti-NSP/ORF) (Y-axis). IgA1 anti-SARS-CoV-2 NSP/ORF = anti-ORF3a, -NSP7. IgA2 anti-SARS-CoV-2 NSP/ORF = anti-ORF3a, -ORF8, -NSP1, -NSP2, ORF4 (E), ORF5 (M). IgG1 anti-SARS-CoV-2 NSP/ORF = anti-ORF3b, -NSP1. IgM anti-SARS-CoV-2 NSP/ORF = anti-ORF8, -NSP1, ORF4 (E). Pearson’s product moment correlation coefficient, p < 0.001.

We also measured host IFN-α2a (IFN-α) cytokine secretion longitudinally in the same matched nasal swab and serum samples. We detected nasal IFN-α at least one week before the peak of anti-IFN-α autoantibodies. While nasal IFN-α peaked as early as one week post-onset, anti-IFN-α autoantibodies continued to increase, reaching peak levels around two weeks post-onset when host IFN-α waned (Fig. 3B). We observed a similar pattern of IFN-α and anti-IFN-α dynamics in the blood (Supplemental Fig. 5). Thus, we show a coordinated balance between IFN-α secretion and subsequent anti-IFN-α production in the nasal mucosa and blood of mild to moderate COVID-19.

Next, we explored whether anti-IFN-α autoantibodies in the nasal mucosa or blood could be associated with symptomatology or other clinical features. When distinguishing between nasal and blood anti-IFN-α producers, we found no differences with regard to sex, age, or BMI (Fig. 3E). We did, however, identify a significant association between anti-IFN-α autoantibodies and disease features, including patient symptoms and biomarkers of systemic inflammation. Patients that developed systemic IgG1 anti-IFN-α had elevated C-reactive protein (CRP), a marker for systemic inflammation (Wilcoxon rank-sum test, p < 0.01) and reported persistent shortness of breath (SOB) and other relevant symptoms (Chi-square test, p < 0.01, Fig. 3E). Of note, SOB is used to indicate disease severity by NIH guidelines and may act as an indication of viral spread to the lower respiratory tract (32). In contrast to systemic IgG1 producers, nasal IgA1 anti-IFN-α autoantibody producers showed significantly lower CRP, SOB, and other symptoms related to disease severity (Fig. 3E). Interestingly, patients who produced IgA2 anti-IFN-α in the absence of IgA1 had elevated CRP levels and even worse clinical symptoms (Fig. 3E), suggesting that IgA2 anti-IFN-α may be induced under exacerbated mucosal hyperinflammatory conditions. Thus, nasal IgA1 anti-IFN-α, but not nasal IgA2 or systemic IgG1, may benefit mild to moderate patients when induced at the mucosal site of infection. Finally, patients that produced nasal IgA1 anti-IFN-α, but not systemic IgG1, showed the highest production of host IFN-α2a in the airways (Fig. 3E), suggesting that anti-IFN-α autoantibodies may be induced as a local feedback loop by the host IFN-α response.

To determine whether the viral-induced anti-IFN-α response was correlated with viral load, we assessed the same nasal swabs for SARS-CoV-2 copy numbers. We found no significant differences in viral load (maximum viral copy numbers/mL by qPCR) between patients that produced either nasal or systemic anti-IFN-α (Fig. 3F). However, we found that anti-IFN-α was secreted days after the peak viral load (Fig. 3F), confirming that nasal anti-IFN-α was induced post-viral-exposure and not pre-existing in our mild to moderate patients. Since nasal anti-IFN-α autoantibodies were associated with efficient disease recovery, we assessed their impact on anti-SARS-CoV-2 antibodies. We found a positive linear correlation between anti-IFN-α and anti-SARS-CoV-2 immunity (Fig. 3G), particularly against non-structural proteins (NSP/ORF), which are known to induce host-cell apoptosis and immunomodulatory functions (6, 33). We found nasal-specific correlations between anti-IFN-α and anti-NSP7/ORF3a and an equivalent blood-specific correlation between anti-IFN-α and anti-NSP1/ORF3b (Fig. 3G). Thus, transient but robust production of nasal IgA1 anti-IFN-α was induced after SARS-CoV-2 infection in patients with mild and moderate COVID-19 and was associated with favorable outcomes, including fewer symptoms and robust anti-SARS-CoV2 immunity.

### Hospitalized patients show elevated anti-IFN-α in the airways associated with uncontrolled IFN-α, higher viral load, and lower anti-SARS-CoV-2 immunity

To determine whether the coordinated balance between anti-IFN-α and IFN-α we identified in mild/moderate-recovered patients is disrupted in severe COVID-19, we performed flowBEAT on endotracheal aspirates (ETA) and peripheral blood from 23 hospitalized and unvaccinated patients admitted to the intensive care unit (ICU) with life-threatening (severe) COVID-19. We detected anti-IFN-α autoantibodies in the airway mucosa (ETA) of 95% of severe patients, while only 43% (10/23) of them had detectable anti-IFN-α in the blood (Fig. 4A). We observed a similar pattern with anti-IFN-ω in the same donors, but at lower levels (Supplemental Fig. 4). In contrast to mild and moderate outpatients, severe patients in the ICU sustained high levels of IFN-α secretion in the ETA that positively correlated with the amount of anti-IFN-α autoantibodies (Fig. 4A), revealing a dysregulated IFN-α hyperinflammatory response at the airway site of infection. Consistent with sustained hyperinflammation in the airways, hospitalized patients produced higher levels of the pro-inflammatory IgA2 anti-IFN-α, unlike mild and moderate patients, which predominantly produced the IgA1 isotype (Fig. 4B). Similarly, the anti-SARS-CoV-2 response (anti-S) and germline autoreactive VH4-34 antibodies, which we previously showed to be increased in the blood of hospitalized patients (34), also switched from the IgA1 to the pro-inflammatory IgA2 isotype in the airways (Fig. 4B).

**Fig. 4:**
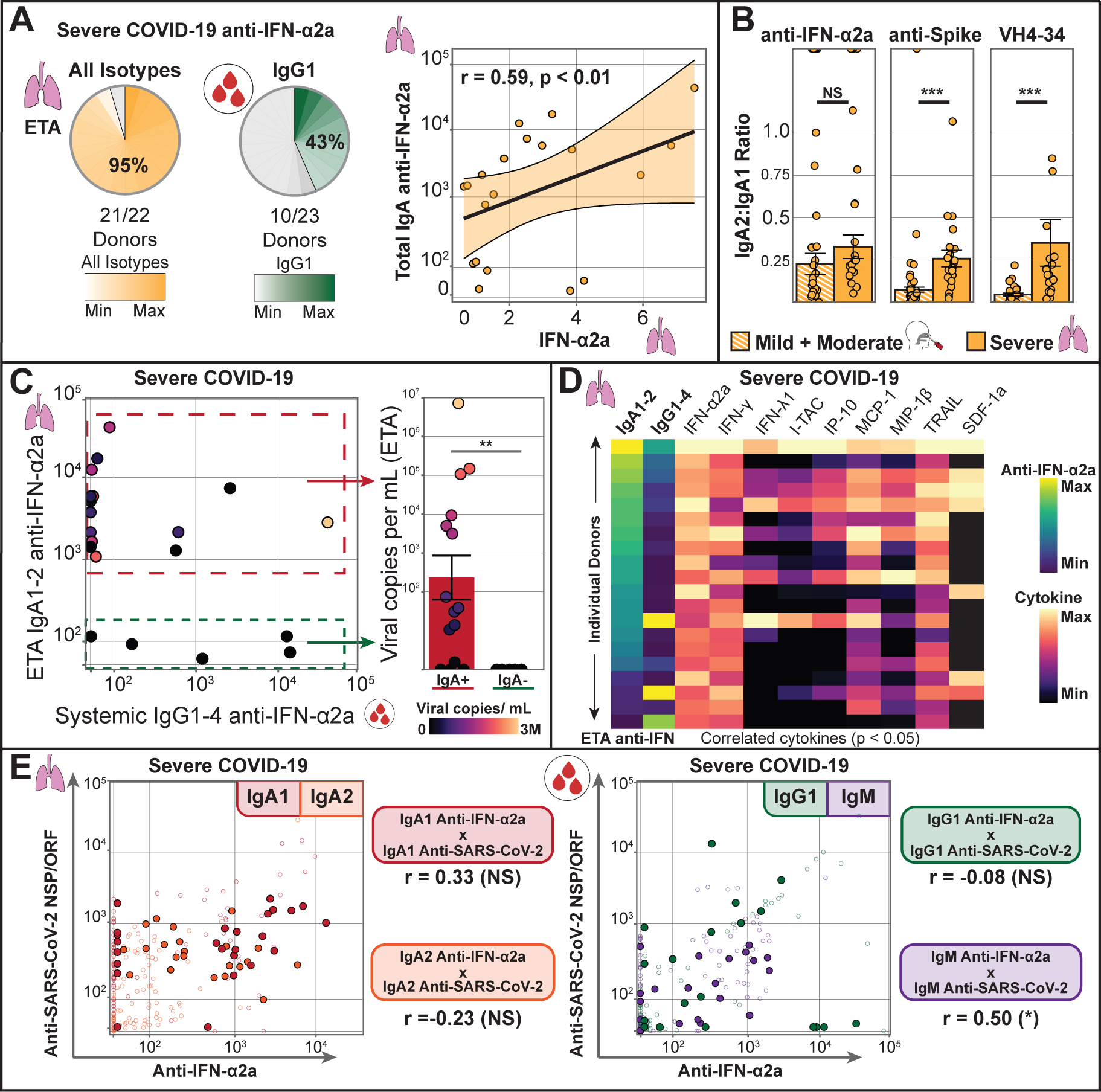
Severe COVID-19 patients show elevated airway anti-IFN-α associated with hyperinflammation and elevated viral load. (A) Pie charts of airway (ETA) and systemic anti-IFN-α2a autoantibody frequency and signal intensity in severe hospitalized COVID-19. The dot plot shows a positive correlation between the ETA’s anti-IFN-α2a autoantibodies (IgA1/2) and IFN-α2a cytokine. Pearson’s product moment correlation coefficient, p < 0.01. (**B**) The bar chart compares the ratio of IgA2:IgA1 isotype usage in the airways of mild/moderate (nasal swabs) and severe (ETA) patients. Wilcoxon rank-sum text, p < 0.001. (**C**) Dot plot of individual hospitalized patients showing systemic IgG(1-4) and airway IgA(1-2) anti-IFN-α2a. Patient populations are gated (dashed lines) as airway IgA-producers (red) and non-producers (green). The bar chart shows viral copies/mL in the ETA of each gated population. (**D**) Heatmap of ETA IgA and IgG anti-IFN-α2a, along with ETA IFN cytokine and other hyper-inflammatory markers correlated to anti-IFN-α2a (Pearson correlation, p<0.05). Rows correspond to individual patients, while columns indicate features as labeled. (**E**) Correlation plots between anti-IFN-α2a (X-axis) and composite anti-non-structural proteins (anti-NSP/ORF) (Y-axis). Colored dots indicate severe COVID-19 and hollow dots indicate mild/moderate COVID-19, as presented in Fig. 3G. Anti-SARS-CoV-2 NSP/ORF programs shown as in Fig. 3G. Pearson’s product moment correlation coefficient, p < 0.05.

Interestingly, airway anti-IFN-α autoantibodies in severe COVID-19 patients were associated with sustained viral load (Wilcoxon rank-sum test, p < 0.01, Fig. 4C), suggesting an ongoing antiviral immune response. Indeed, airway anti-IFN-α correlated positively with increased inflammatory cytokines along with elevated signals of myeloid recruitment and activation, including IFN-γ, IFN-λ1, I-TAC, IP-10, MCP-1, MIP-1β, TRAIL, and SDF-1a (Fig. 4D). However, while anti-IFN-α in mild and moderate patients correlated with enhanced anti-SARS-CoV-2 immunity (i.e., increased anti-NSP, - ORF, -M, and -E in the airway and blood, Fig. 3F), this positive correlation no longer persisted in the airways of severe patients (Fig. 4E). Thus, in hospitalized patients, higher levels of airway IgA2 anti-IFN-α in the absence of IgA1 or the presence of high systemic IgG1 correlated with higher viral load, hyperinflammation, lower anti-SARS-CoV-2 immunity and a more severe disease course.

## DISCUSSION

The epithelium of the nasal mucosa is the initial site of SARS-CoV-2 exposure and infection. As such, antiviral immune responses mounted locally in the nasal mucosa can play a role in determining disease recovery or progression to severe COVID-19. Here, we show that transient autoantibodies against type-I IFN (anti-IFN-α) in the nasal mucosa are a common SARS-CoV-2-induced immune response associated with efficient recovery from COVID-19 disease. Specifically, mild and moderate-recovered patients who produced nasal, but not systemic, IgA1 anti-IFN-α early during infection showed lesser symptoms and increased anti-SARS-CoV-2 immunity in the airways. In contrast, if the initial nasal-specific anti-IFN-α progressed to a systemic IgG1 response in the blood, it was associated with disease severity, including worsened symptoms and increased systemic inflammation, corroborated by a switch in anti-SARS-CoV-2 antibody subclass from IgG1 to a more inflammatory IgG3. Thus, the high prevalence (> 70%) of nasal anti-IFN-α in mild/moderate-recovered patients suggests that autoreactivity to type-I IFN in the airways is an intrinsic aspect of the host protective antiviral immunity and not a pre-condition to severe COVID-19, as previously suggested for pre-existing blood anti-IFN-α (16). Most importantly, our findings on the potential protective role of anti-type I IFN in disease resolution should warn against the consequences of either depleting anti-IFN-α or administering nasal type-I IFN in COVID-19 patients, both of which are ongoing clinical trials (35, 36).

Type-I IFN is an essential antiviral cytokine that protects against most viral infections (6). However, its role in efficient recovery from SARS-CoV-2 and other coronavirus infections is uncertain. Animal models of coronavirus infection in mice (9, 10) and non-human primates (NHP) (11) suggest that type-I IFN is dispensable or detrimental to resolving the disease. Mice that lack the IFN-αβ receptor (IFNAR-/-) are protected from lethal infection with SARS-CoV without affecting viral load (9). Similarly, a mouse model of SARS-CoV-2 infection shows that type-I IFN is not required for viral clearance but instead is responsible for the airway infiltration of inflammatory cells and drives the lung pathology associated with severe disease (10). In NHP, persistent type-I IFN can lead to lung pathology but could be protective if produced in small amounts and only at the early stages of the SARS-CoV-2 infection (11). Thus, our data corroborate animal models and support a mechanism in which the virus induces transient anti-IFN-α autoantibodies early during infection, perhaps to counteract the detrimental effects of otherwise excessive or persistent IFN-α production. As the disease progresses from mild/moderate to severe in hospitalized ICU patients, the protective IFN-α/anti-IFN-α axis is dysregulated, resulting in uncontrolled and persistent IFN-α and anti-IFN-α production in the airways and blood.

Previous studies showed that blood anti-IFN-α and inborn errors of type-I IFN immunity are associated with increased susceptibility to severe, life-threatening COVID-19 (12, 16). Conclusions drawn from these previous works infer that anti-IFN-α is a pre-existing autoantibody, supported by their findings showing no detectable anti-IFN-α in the blood of mild patients measured at disease onset, while systemic anti-IFN-α was detected in over 10% of hospitalized COVID-19 patients. These findings would suggest that pre-existing, instead of viral-induced, systemic anti-IFN-α predisposes patients to severe disease and hospitalization. However, these previous studies measured anti-IFN-α in the blood, not airways, early after disease onset. To reconcile the apparent discrepancy to our findings, we show that mild patients develop systemic anti-IFN-α only later, more than two weeks post-disease onset. Thus, it is possible that previous studies could have missed the viral-induced anti-IFN-α in mild patients as they were measured in blood early in disease onset when anti-IFN-α is only detectable in the airways. Similarly, some hospitalized patients who showed systemic anti-IFN-α in previous studies may have developed their autoantibodies after a mild or moderate SARS-CoV-2 infection and before progressing to severe COVID-19 and hospitalization. Indeed, a separate study corroborates our findings supporting viral-induced anti-IFN-α response by identifying new-onset anti-IFN-α in the blood of severe COVID-19 (37). Furthermore, while some studies suggest that autoimmune polyendocrine syndrome type-1 (APS-1) patients with pre-existing systemic anti-IFN-α are more susceptible to severe COVID-19 (38), another prospective study analyzing APS-1 patients with high titers of pre-existing systemic anti-IFN-α showed only mild symptoms of COVID-19 (31). Altogether, we and others confirm that anti-IFN-α can be induced by SARS-CoV-2 but do not necessarily lead to life-threatening COVID-19 disease.

The prevalent induction of anti-IFN-α autoantibodies in the nasal mucosa of mild/moderate (> 70%) and severely-infected (> 95%) COVID-19 patients reveal that autoreactive B-cell clones against IFN-α are ubiquitously present in the population. Indeed, recent studies show that a broad new-onset autoantibody response against inflammatory cytokines and chemokines is a common feature of the immune response against COVID-19 (17, 37, 39), and their neutralizing capacity likely plays a role in attenuating hyperinflammatory responses during infection (40). Notably, these autoantibodies were not associated with increased disease severity but with an efficient resolution, similar to our findings for airway anti-IFN-α. Thus, producing autoantibodies against inflammatory cytokines might represent a common immune mechanism to control or resolve hyperinflammation during viral infections.

The mechanisms that lead to the activation of autoreactive B cells against cytokines, particularly IFN-α, are unknown. Our data, particularly the positive association between IFN-α levels and subsequent anti-INF-α production, suggest that activation of autoreactive B cells is antigen-specific and dependent on IFN-α. All patients that produced IFN-α in the airways developed anti-IFN-α, which waned completely when IFN-α was no longer detected. Since the kinetics of anti-IFN-α in the airways follows IFN-α secretion and since severe patients that sustain high levels of IFN-α had corresponding higher anti-IFN-α, autoreactive B cells are likely activated locally by IFN-α via their B-cell receptor (BCR). Indeed, patients who did not secrete detectable levels of IFN-α in the airways did not produce anti-IFN-α. Moreover, the quick and transient nature of the airway autoantibodies, which wane with disease recovery, suggests that B cells are quickly activated independent of germinal centers or T-cell help, leading to short-lived (transient) antibody-secreting cells (41, 42). In contrast, when analyzing the blood, we found anti-IFN-α autoantibodies that persisted longer than 2 months, suggesting a dysregulated systemic B-cell activation and generation of long-lived antibody-secreting cells in a small group of potentially at-risk patients.

Finally, viral-induced anti-IFN-α in blood without airway anti-IFN-α characterizes patients with a worse prognosis and may carry implications for future viral defense. If measured outside the context of immediate SARS-CoV-2 infection and after disease recovery, the persistent blood anti-IFN-α could be interpreted as “pre-existing” (as previously suggested (16, 30)) and identify patients at potential risk of re-infections or post-acute sequelae of COVID-19 (PASC) and long-COVID symptoms. Thus, an early and transient nasal IgA1 anti-IFN-α without progression to blood IgG1 anti-IFN-α is a predictive biomarker of protection and prognosis for efficient recovery from COVID-19.

Altogether, our findings challenge the current notion that anti-IFN-α autoantibodies are a pathologic feature of COVID-19 and instead support a model in which nasal anti-IFN-α may serve an important regulatory function to counteract the detrimental effects of excessive IFN-α and restore homeostasis following SARS-CoV-2 invasion of the respiratory mucosa. Further, our data also challenge the notion that systemic anti-IFN-α is a risk factor for life-threatening COVID-19. Instead, we show that the progression from a localized nasal IgA1 anti-IFN-α to a systemic IgG1 anti-IFN-α is a sign of uncontrolled disease and a biomarker of disease severity or worse prognosis in all patient groups. Future studies are needed to determine the opposing roles of airway versus blood anti-IFN-α in COVID-19 and whether persistent viral-induced anti-IFN-α in the blood can predispose to subsequent viral infections and PASC. Moreover, subsequent studies should determine whether nasal anti-IFN-α autoantibody response is a common protection mechanism against other respiratory viral infections, including Influenza and viruses that infect other mucosal sites.

## METHODS

### Study participants and ethical approvals

The study included 129 participants enrolled through the University of California San Francisco (UCSF)’s COVID-19 Host Immune Response Pathogenesis (CHIRP) study, Emory University’s PROATECT study previously described (1), and the pre-pandemic and healthy control samples were provided by the Clinical and Translational Discovery Core at Children’s Healthcare of Atlanta (CHOA) and Emory University under IRB 00089506. All participants were assigned a new, sequential ID in the format “ERxxx.” IDs are known only to the study investigators as per Emory IRB 00003368.

*CHIRP study*: eligible participants were identified by confirmed SARS-CoV-2 RT-PCR nasal swab test results and had matched plasma and nasal swab biospecimens collected between 4/2020 – 11/2022 (Supplemental Tables 2, 3). All participants provided written informed consent. Longitudinal biospecimens were collected at weeks 0, 1, 3, 10, and 24 from baseline visits. A subset of participants also provided additional samples post-vaccination (2 weeks post-dose #1, 2 weeks post-dose #2, 8 weeks post-dose #1 and 2 weeks post-dose #4). Participants completed a detailed questionnaire at each study visit, including demographic, social, medical history, concomitant medication use, and symptom severity reporting as previously described (43) (Supplemental Table 2). The date of symptom onset (or lack of symptoms, if asymptomatic disease) was recorded at the baseline visit. COVID-19 symptom severity (mild versus moderate) was defined based on symptom burden (total number of reported symptom categories) and duration (weeks of symptoms) (Supplemental Table 2). Clinical labs, including SARS-CoV-2 viral load (nasal swab), complete blood count with differential, comprehensive metabolic panel, erythrocyte sedimentation rate (ESR), and high sensitivity C-reactive protein (hs-CRP) were collected at each visit, as well as whole blood for processing for plasma and serum. This study was approved by the UCSF and Emory University Committee on Human Subjects Research (UCSF IRB 20-30588; Emory IRB 00003368. All participants provided written informed consent, and all biospecimens used for research were de-identified.

*Emory PROATECT study*: eligible participants were identified by confirmed SARS-CoV-2 RT-PCR test results and enrolled as previously described (1) (Supplemental Table 3). Briefly, 23 patients with severe, life-threatening COVID-19 were recruited from the ICU of Emory University, Emory St. Joseph’s, Emory Decatur, and Emory Midtown Hospitals between 6/2020 – 2/2021. We also recruited 24 outpatients with mild and moderate COVID-19 in the Emory Acute Respiratory Clinic. Outpatients were classified by disease severity (mild or moderate) following NIH guidelines (32) using symptom data collected by a healthcare provider (Supplemental Table 3). Infection-naïve patients were recruited for pre- and post-vaccination antibody assessments in blood and nasal swabs. Informed consent was obtained for all study participants. Emory’s IRB approved all studies under protocol numbers 00003368, 00058507, 00057983, and 00058271.

### Sample processing

*Plasma and serum*: samples were collected for all study participants, regardless of COVID-19 diagnosis or severity. To obtain serum, whole peripheral blood was collected and allowed to clot, and supernatant serum was isolated and cryopreserved. For plasma collection, whole blood was collected in EDTA tubes, and plasma was isolated by centrifugation at 400 × g for 10 minutes at 4°C followed by removal of the supernatant. To further precipitate platelets, contaminating cells, or remaining debris, the isolated plasma and serum were centrifuged at 4000 × g for 10 minutes at 4°C. Samples were distributed in aliquots and cryopreserved at -80C.

*Nasal swabs:* samples were collected from both nostrils and transferred to 3 mL of viral preservation media. Swabs (in media) were vortexed to liberate collected antibodies into media. Samples were further mechanically dissociated using a syringe and 25 Ga needle to disrupt aggregates of mucins and other respiratory secretions. Samples were centrifuged at 2000 × g to precipitate non-soluble aggregates. Processing of samples from SARS-CoV-2 positive donors was performed within a biosafety cabinet following BSL2+ protocols approved by the Emory University Institutional Biosafety Committee and Biosafety Officer.

*Endotracheal aspirates (ETA)*: samples were collected by healthcare providers in the ICU from either endotracheal intubation or tracheostomy (Supplemental Table 5). ETA samples were transferred to a USDA-approved BSL3 containment facility at Emory for further processing. To disrupt mucin aggregates, ETA was mixed 1:1 with a 50 mM EDTA solution (final concentration 25 mM EDTA) in custom RPMI-1640 media deficient in biotin, L-glutamine, phenol red, riboflavin, and sodium bicarbonate (defRPMI-1640) and dissociated using a syringe as previously described (1). Samples were centrifuged at 250 × g, and supernatants were cryopreserved at -80°C. Before removal from BSL3 containment, ETA supernatants were UV-inactivated following our published protocols (44).

### Antigen preparation and conjugation

All SARS-CoV-2 and auto-antigens were obtained from commercial sources, except for SARS-CoV-2 non-structural antigens, which were produced in collaboration with ChemPartner (Supplemental Table 1). All antigens, including a purified bovine serum albumin (BSA) control antigen, were biotin-conjugated using EZ-link NHS-biotin reagents at a 20-fold molar excess. Antigens were loaded onto commercially sourced streptavidin-coated beads at an empirically determined saturation point in 50 µL of PBS + 0.05% BSA + 0.2% Tween-20 (PBS-BSA) and allowed to bind for 20 minutes on ice before the excess (uncaptured) antigen was removed by washing with additional PBS-BSA.

### Fluorescent detection antibodies

Fluorescently conjugated monoclonal antibodies were obtained from commercial sources for the specific detection of eight human antibody isotypes and subclasses (IgG1, IgG2, IgG3, IgG4, IgA1, IgA2, IgE, and IgM). IgG1-, IgA2-, IgE- and IgM-specific antibodies were purchased with fluorophores pre-conjugated by Southern Biotech, while IgG2-, IgG3-, IgG4- and IgA1-specific antibodies were conjugated using the Lightning-Link fluorescent conjugation kits (Abcam) following the optimal manufacturer protocol. We used mouse monoclonal anti-SARS-CoV-2 and anti-interferon antibodies to determine assay sensitivity and specificity, and a secondary goat polyclonal anti-mouse (PE or AF488 fluorophore) to reveal monoclonal anti-antigen captured on each bead species.

### FlowBEAT calibration

We experimentally determined the antigen-bead saturation point for RBD, S1, S2, IFN-α2a, and IFN-ω antigens. All were found to have an identical saturation point of 2 pmol per 50,000 beads in 50 µL of PBS-BSA. We then confirmed the capacity of saturated beads to capture antigen-specific antibodies by flow cytometry. To confirm the antigen specificity signal for each mouse monoclonal antibody, we performed serial dilutions of antigen-specific antibodies co-incubated with antigen-saturated beads in PBS-BSA and revealed by a secondary stain with fluorescent anti-mouse. Each antigen-specific antibody had a maximum signal output at least 1000-fold greater than the unstained control, which decreases linearly with serial dilution.

### FlowBEAT assays

After thawing, samples were centrifuged at 2,000 × g for 10 minutes at 4°C to precipitate any remaining aggregates. Serum or plasma samples were diluted to a final concentration of 1:125 with PBS-BSA in a reaction mix containing beads coated with antigen at an empirically determined saturation point. Airway samples were diluted to a final concentration of 1:2. Samples and beads were incubated on ice in the dark for 30 minutes, mixing at least every 15 minutes to prevent beads from settling. Beads were washed with PBS-BSA and resuspended before fluorescent antibody staining. Antibody staining was performed by mixing equal volumes of resuspended beads and a master mix containing eight fluorescently conjugated monoclonal antibodies specific to IgG1, IgG2, IgG3, IgG4, IgA1, IgA2, IgE and IgM. The optimal concentration of each antibody species was empirically determined by titration. The reaction was incubated on ice in the dark for 30 minutes, mixing at least every 15 minutes. Stained beads were washed with PBS-BSA and resuspended in 1X PBS. Samples were fixed in 1:6 final dilution of FACS Lysing solution (BD Biosciences), which we showed inactivates SARS-CoV-2 (44). While fixing, samples were kept in the dark at room temperature for 20 minutes, mixing at least every 10 minutes. A final wash with PBS-BSA was performed, and beads were resuspended in PBS-BSA. All samples were analyzed within 24 hours of staining using a 5-laser Cytek Aurora flow cytometer. All data collection occurred on the same instrument, and the cytometer was calibrated with a consistent lot of Cytek SpectroFlo QC beads before each data collection. To standardize each flowBEAT batch, we established a daily standard plasma sample with strong reactivity to SARS-CoV-2 spike subunits. The standard positive and a sample-free control were included in all flowBEAT assays to confirm the assay’s performance and control for potential variations.

### Cytokine quantification

The levels of human cytokines in serum, nasal swabs, and ETA were measured using a Meso Scale Discovery U-plex custom multiplex assay kit, as we previously described (1). We measured an additional ten analytes (IFN-α2a, IFN-γ, IL-15, IL-29/IFN-λ1, IP-10, MCP-1, MDC, MIP-3β, SDF-1a, and TRAIL) following the manufacturer’s protocol (Meso Scale Discovery) in plasma, nasal swabs, and ETA, which were prepared following our established UVC-inactivation protocol (44).

### Computational analysis

Flow Cytometry Standard (FCS) files were gated and analyzed in FlowJo version 10 software (BD Biosciences) to identify median fluorescence intensity (MFI) values for each antigen-specific antibody isotype fluorescence (example gating strategy in Supplementary Fig. 1). Exported MFI values were background-corrected using a linear regression correlating background antigen-specific signal (as determined in pre-pandemic donors) and control BSA signal. Figures were generated using R version 4.2.2 using the packages “ggplot2”, “ggcyto,” and “flowcore.” The code will be available upon publication on the Ghosn Lab’s GitHub page: github.com/Ghosn-Lab.

## Data Availability

All data produced in the present study are available upon reasonable request to the authors. The code will be available on the Ghosn Lab GitHub page: github.com/Ghosn-Lab.

## ACKNOWLEDGEMENTS

We thank all the patients and their families for participating in this study. We also thank Chunhua Liu, Junhui Zhang, Yuliang Dong, Feimin Lin, Cheng Fang and Greg Liang (ChemPartner) for producing and purifying SARS-CoV-2 proteins; David E. Gordon and Nevan J. Krogan (Gladstone Institutes and University of California, San Francisco) for providing SARS-CoV-2 expression constructs; Sang N. Le, John Varghese, Anum Jalal, Saeyun Lee and Rahul Patel (Emory University) for patient recruitment; the nurses, staff, and providers in the 71 ICU at Emory University Hospital Midtown, the medical ICU in Emory Decatur Hospital, the 5G/6G ICU in Emory University Hospital, and the ICU in Emory Saint Joseph’s Hospital for caring for patients enrolled in this study; the Emory Biosafety Officers Kalpana Rengarajan and Esmeralda Meyer for their assistance in establishing biosafety protocols and facilities. Research reported in this publication was supported in part by the Emory Pediatrics/Winship Flow Cytometry Core, by the Children’s Healthcare of Atlanta (CHOA) and Emory University’s Clinical and Translational Discovery Core (CCTDC), and by the Emory Multiplexed Immunoassay Core (EMIC).

## Notes

### Competing Interest Statement

The authors have declared no competing interest.

### Funding Statement

This study was funded by the National Institutes of Health (NIH) National Institute of Allergy and Infectious Diseases (NIAID) awards R21AI167032 (E.E.B.G.) and R01AI123126-05S1 (E.E.B.G.); and by the Program for Breakthrough Biomedical Research Award (N.R.R., S.A.L., E.E.B.G.).

### Author Declarations

The Institutional Review Board (IRB) of Emory University gave ethical approval for this work under IRB protocol #00003368

## REFERENCES

1. Eddins DJ, Yang J, Kosters A, Giacalone VD, Pechuan-Jorge X, Chandler JD, et al. Transcriptional reprogramming of infiltrating neutrophils drives lung pathology in severe COVID-19 despite low viral load. Blood Adv. 2023;7(5):778–99.

2. Bergamaschi L, Mescia F, Turner L, Hanson AL, Kotagiri P, Dunmore BJ, et al. Longitudinal analysis reveals that delayed bystander CD8+ T cell activation and early immune pathology distinguish severe COVID-19 from mild disease. Immunity. 2021;54(6):1257–75 e8.

3. Lucas C, Wong P, Klein J, Castro TBR, Silva J, Sundaram M, et al. Longitudinal analyses reveal immunological misfiring in severe COVID-19. Nature. 2020;584(7821):463-9.

4. Dobosh B, Zandi K, Giraldo DM, Goh SL, Musall K, Aldeco M, et al. Baricitinib attenuates the proinflammatory phase of COVID-19 driven by lung-infiltrating monocytes. Cell Rep. 2022;39(11):110945.

5. Hadjadj J, Yatim N, Barnabei L, Corneau A, Boussier J, Smith N, et al. Impaired type I interferon activity and inflammatory responses in severe COVID-19 patients. Science. 2020;369(6504):718-24.

6. Park A, Iwasaki A. Type I and Type III Interferons - Induction, Signaling, Evasion, and Application to Combat COVID-19. Cell Host Microbe. 2020;27(6):870-8.

7. Galbraith MD, Kinning KT, Sullivan KD, Araya P, Smith KP, Granrath RE, et al. Specialized interferon action in COVID-19. Proc Natl Acad Sci U S A. 2022;119(11).

8. Sposito B, Broggi A, Pandolfi L, Crotta S, Clementi N, Ferrarese R, et al. The interferon landscape along the respiratory tract impacts the severity of COVID-19. Cell. 2021;184(19):4953–68 e16.

9. Channappanavar R, Fehr AR, Vijay R, Mack M, Zhao J, Meyerholz DK, et al. Dysregulated Type I Interferon and Inflammatory Monocyte-Macrophage Responses Cause Lethal Pneumonia in SARS-CoV-Infected Mice. Cell Host Microbe. 2016;19(2):181–93.

10. Israelow B, Song E, Mao T, Lu P, Meir A, Liu F, et al. Mouse model of SARS-CoV-2 reveals inflammatory role of type I interferon signaling. J Exp Med. 2020;217(12).

11. Viox EG, Hoang TN, Upadhyay AA, Nchioua R, Hirschenberger M, Strongin Z, et al. Modulation of type I interferon responses potently inhibits SARS-CoV-2 replication and inflammation in rhesus macaques. Sci Immunol. 2023;8(85):eadg0033.

12. Zhang Q, Bastard P, Liu Z, Le Pen J, Moncada-Velez M, Chen J, et al. Inborn errors of type I IFN immunity in patients with life-threatening COVID-19. Science. 2020;370(6515).

13. Pairo-Castineira E, Clohisey S, Klaric L, Bretherick AD, Rawlik K, Pasko D, et al. Genetic mechanisms of critical illness in COVID-19. Nature. 2021;591(7848):92-8.

14. Liechti T, Iftikhar Y, Mangino M, Beddall M, Goss CW, O’Halloran JA, et al. Immune phenotypes that are associated with subsequent COVID-19 severity inferred from post-recovery samples. Nat Commun. 2022;13(1):7255.

15. van der Wijst MGP, Vazquez SE, Hartoularos GC, Bastard P, Grant T, Bueno R, et al. Type I interferon autoantibodies are associated with systemic immune alterations in patients with COVID-19. Sci Transl Med. 2021;13(612):eabh2624.

16. Bastard P, Gervais A, Le Voyer T, Rosain J, Philippot Q, Manry J, et al. Autoantibodies neutralizing type I IFNs are present in ∼4% of uninfected individuals over 70 years old and account for ∼20% of COVID-19 deaths. Sci Immunol. 2021;6(62).

17. Wang EY, Mao T, Klein J, Dai Y, Huck JD, Jaycox JR, et al. Diverse functional autoantibodies in patients with COVID-19. Nature. 2021;595(7866):283-8.

18. Combes AJ, Courau T, Kuhn NF, Hu KH, Ray A, Chen WS, et al. Global absence and targeting of protective immune states in severe COVID-19. Nature. 2021;591(7848):124-30.

19. Vazquez SE, Bastard P, Kelly K, Gervais A, Norris PJ, Dumont LJ, et al. Neutralizing Autoantibodies to Type I Interferons in COVID-19 Convalescent Donor Plasma. J Clin Immunol. 2021;41(6):1169–71.

20. Lee JS, Park S, Jeong HW, Ahn JY, Choi SJ, Lee H, et al. Immunophenotyping of COVID-19 and influenza highlights the role of type I interferons in development of severe COVID-19. Sci Immunol. 2020;5(49).

21. Broggi A, Ghosh S, Sposito B, Spreafico R, Balzarini F, Lo Cascio A, et al. Type III interferons disrupt the lung epithelial barrier upon viral recognition. Science. 2020;369(6504):706-12.

22. Hatton CF, Botting RA, Duenas ME, Haq IJ, Verdon B, Thompson BJ, et al. Delayed induction of type I and III interferons mediates nasal epithelial cell permissiveness to SARS-CoV-2. Nat Commun. 2021;12(1):7092.

23. Major J, Crotta S, Llorian M, McCabe TM, Gad HH, Priestnall SL, et al. Type I and III interferons disrupt lung epithelial repair during recovery from viral infection. Science. 2020;369(6504):712-7.

24. Center SC. Human SARS-COV-2 Serology Standard: Frederick National Laboratory for Cancer Research; 2023 [Available from: https://frederick.cancer.gov/initiatives/seronet/serology-standard.

25. Murray SM, Ansari AM, Frater J, Klenerman P, Dunachie S, Barnes E, et al. The impact of pre-existing cross-reactive immunity on SARS-CoV-2 infection and vaccine responses. Nat Rev Immunol. 2023;23(5):304–16.

26. Focosi D, Maggi F, Casadevall A. Mucosal Vaccines, Sterilizing Immunity, and the Future of SARS-CoV-2 Virulence. Viruses. 2022;14(2).

27. Mes L, Steffen U, Chen HJ, Veth J, Hoepel W, Griffith GR, et al. IgA2 immune complexes selectively promote inflammation by human CD103(+) dendritic cells. Front Immunol. 2023;14:1116435.

28. Steffen U, Koeleman CA, Sokolova MV, Bang H, Kleyer A, Rech J, et al. IgA subclasses have different effector functions associated with distinct glycosylation profiles. Nat Commun. 2020;11(1):120.

29. Damelang T, Rogerson SJ, Kent SJ, Chung AW. Role of IgG3 in Infectious Diseases. Trends Immunol. 2019;40(3):197–211.

30. Bastard P, Rosen LB, Zhang Q, Michailidis E, Hoffmann HH, Zhang Y, et al. Autoantibodies against type I IFNs in patients with life-threatening COVID-19. Science. 2020;370(6515).

31. Meisel C, Akbil B, Meyer T, Lankes E, Corman VM, Staudacher O, et al. Mild COVID-19 despite autoantibodies against type I IFNs in autoimmune polyendocrine syndrome type 1. J Clin Invest. 2021;131(14).

32. Coronavirus Disease 2019 (COVID-19) Treatment Guidelines https://www.covid19treatmentguidelines.nih.gov/: National Institutes of Health; 2023 [updated March 6, 2023.

33. Li Y, Xu Z, Lei Q, Lai DY, Hou H, Jiang HW, et al. Antibody landscape against SARS-CoV-2 reveals significant differences between non-structural/accessory and structural proteins. Cell Rep. 2021;36(2):109391.

34. Woodruff MC, Ramonell RP, Haddad NS, Anam FA, Rudolph ME, Walker TA, et al. Dysregulated naive B cells and de novo autoreactivity in severe COVID-19. Nature. 2022;611(7934):139-47.

35. Aurélien M. Plasma Exchange in Covid-19 Patients With Anti-interferon Autoantibodies clinicaltrials.gov 2022 [Available from: https://clinicaltrials.gov/study/NCT05182515?cond=Interferon%20covid-19&locStr=France&country=France&distance=50&rank=3#collaborators-and-investigators.

36. Lanoix J-P. Treatment of COVID-19 by Nebulization of Inteferon Beta 1b Efficiency and Safety Study clinicaltrials.gov 2023 [Available from: https://clinicaltrials.gov/study/NCT04469491?cond=Interferon%20covid-19&rank=1#collaborators-and-investigators.

37. Chang SE, Feng A, Meng W, Apostolidis SA, Mack E, Artandi M, et al. New-onset IgG autoantibodies in hospitalized patients with COVID-19. Nat Commun. 2021;12(1):5417.

38. Bastard P, Orlova E, Sozaeva L, Levy R, James A, Schmitt MM, et al. Preexisting autoantibodies to type I IFNs underlie critical COVID-19 pneumonia in patients with APS-1. J Exp Med. 2021;218(7).

39. Bodansky A, Wang CY, Saxena A, Mitchell A, Kung AF, Takahashi S, et al. Autoantigen profiling reveals a shared post-COVID signature in fully recovered and long COVID patients. JCI Insight. 2023;8(11).

40. Muri J, Cecchinato V, Cavalli A, Shanbhag AA, Matkovic M, Biggiogero M, et al. Autoantibodies against chemokines post-SARS-CoV-2 infection correlate with disease course. Nat Immunol. 2023;24(4):604–11.

41. Jenks SA, Cashman KS, Woodruff MC, Lee FE, Sanz I. Extrafollicular responses in humans and SLE. Immunol Rev. 2019;288(1):136–48.

42. Woodruff MC, Ramonell RP, Nguyen DC, Cashman KS, Saini AS, Haddad NS, et al. Extrafollicular B cell responses correlate with neutralizing antibodies and morbidity in COVID-19. Nat Immunol. 2020;21(12):1506–16.

43. Peluso MJ, Kelly JD, Lu S, Goldberg SA, Davidson MC, Mathur S, et al. Persistence, Magnitude, and Patterns of Postacute Symptoms and Quality of Life Following Onset of SARS-CoV-2 Infection: Cohort Description and Approaches for Measurement. Open Forum Infect Dis. 2022;9(2):ofab640.

44. Eddins DJ, Bassit LC, Chandler JD, Haddad NS, Musall KL, Yang J, et al. Inactivation of SARS-CoV-2 and COVID-19 Patient Samples for Contemporary Immunology and Metabolomics Studies. Immunohorizons. 2022;6(2):144–55.

