## Supplemental Tables 2-5 for "Transient anti-interferon autoantibodies in the airways are associated with efficient recovery from COVID-19"

### Slide 1
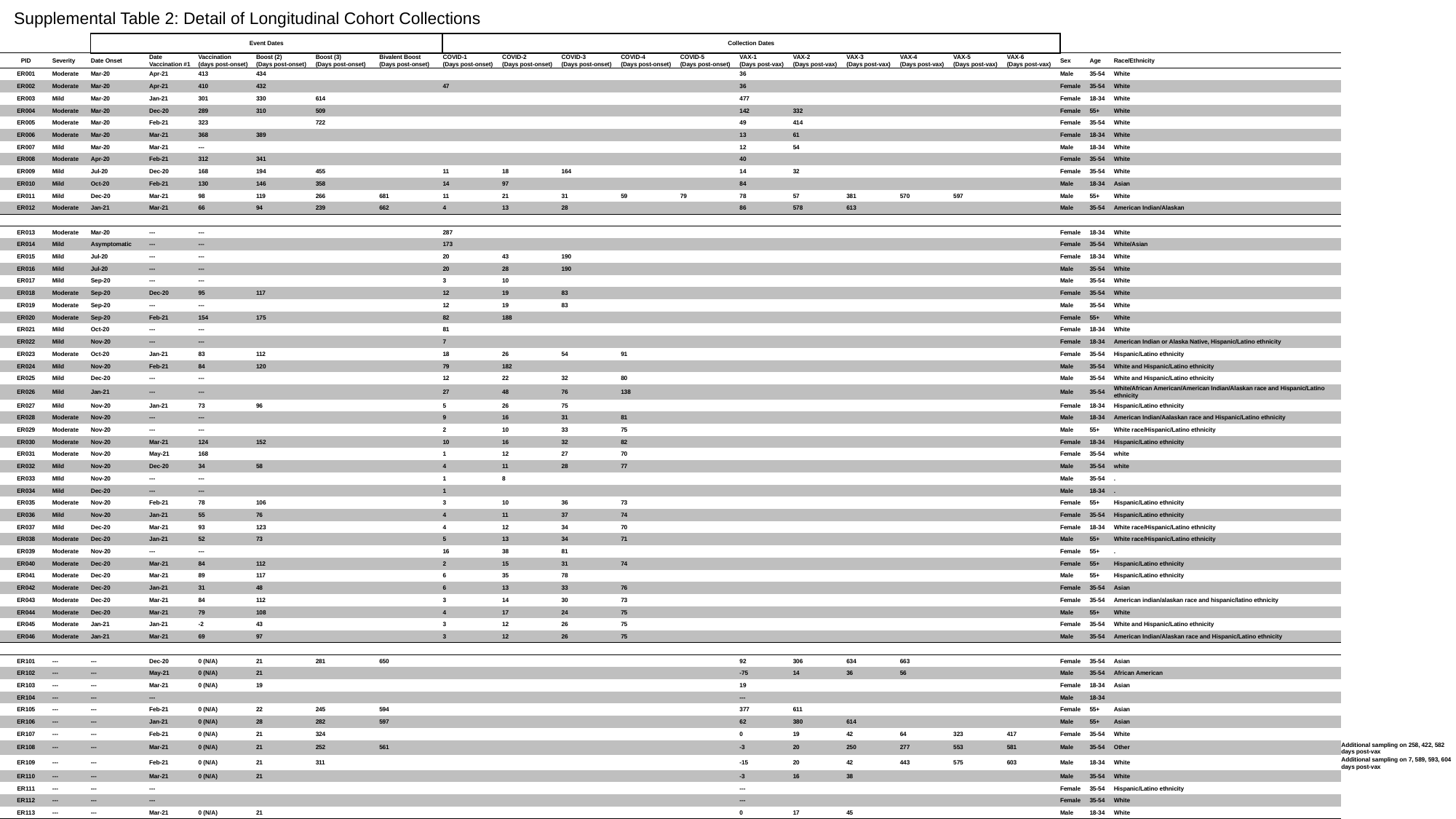

Supplemental Table 2: Detail of Longitudinal Cohort Collections
| | | Event Dates | | | | | | Collection Dates | | | | | | | | | | | | | | |
| --- | --- | --- | --- | --- | --- | --- | --- | --- | --- | --- | --- | --- | --- | --- | --- | --- | --- | --- | --- | --- | --- | --- |
| PID | Severity | Date Onset | Date Vaccination #1 | Vaccination (days post-onset) | Boost (2) (Days post-onset) | Boost (3) (Days post-onset) | Bivalent Boost (Days post-onset) | COVID-1 (Days post-onset) | COVID-2 (Days post-onset) | COVID-3 (Days post-onset) | COVID-4 (Days post-onset) | COVID-5 (Days post-onset) | VAX-1 (Days post-vax) | VAX-2 (Days post-vax) | VAX-3 (Days post-vax) | VAX-4 (Days post-vax) | VAX-5 (Days post-vax) | VAX-6 (Days post-vax) | Sex | Age | Race/Ethnicity | |
| ER001 | Moderate | Mar-20 | Apr-21 | 413 | 434 | | | | | | | | 36 | | | | | | Male | 35-54 | White | |
| ER002 | Moderate | Mar-20 | Apr-21 | 410 | 432 | | | 47 | | | | | 36 | | | | | | Female | 35-54 | White | |
| ER003 | Mild | Mar-20 | Jan-21 | 301 | 330 | 614 | | | | | | | 477 | | | | | | Female | 18-34 | White | |
| ER004 | Moderate | Mar-20 | Dec-20 | 289 | 310 | 509 | | | | | | | 142 | 332 | | | | | Female | 55+ | White | |
| ER005 | Moderate | Mar-20 | Feb-21 | 323 | | 722 | | | | | | | 49 | 414 | | | | | Female | 35-54 | White | |
| ER006 | Moderate | Mar-20 | Mar-21 | 368 | 389 | | | | | | | | 13 | 61 | | | | | Female | 18-34 | White | |
| ER007 | Mild | Mar-20 | Mar-21 | --- | | | | | | | | | 12 | 54 | | | | | Male | 18-34 | White | |
| ER008 | Moderate | Apr-20 | Feb-21 | 312 | 341 | | | | | | | | 40 | | | | | | Female | 35-54 | White | |
| ER009 | Mild | Jul-20 | Dec-20 | 168 | 194 | 455 | | 11 | 18 | 164 | | | 14 | 32 | | | | | Female | 35-54 | White | |
| ER010 | Mild | Oct-20 | Feb-21 | 130 | 146 | 358 | | 14 | 97 | | | | 84 | | | | | | Male | 18-34 | Asian | |
| ER011 | Mild | Dec-20 | Mar-21 | 98 | 119 | 266 | 681 | 11 | 21 | 31 | 59 | 79 | 78 | 57 | 381 | 570 | 597 | | Male | 55+ | White | |
| ER012 | Moderate | Jan-21 | Mar-21 | 66 | 94 | 239 | 662 | 4 | 13 | 28 | | | 86 | 578 | 613 | | | | Male | 35-54 | American Indian/Alaskan | |
| ER013 | Moderate | Mar-20 | --- | --- | | | | 287 | | | | | | | | | | | Female | 18-34 | White | |
| ER014 | Mild | Asymptomatic | --- | --- | | | | 173 | | | | | | | | | | | Female | 35-54 | White/Asian | |
| ER015 | Mild | Jul-20 | --- | --- | | | | 20 | 43 | 190 | | | | | | | | | Female | 18-34 | White | |
| ER016 | Mild | Jul-20 | --- | --- | | | | 20 | 28 | 190 | | | | | | | | | Male | 35-54 | White | |
| ER017 | Mild | Sep-20 | --- | --- | | | | 3 | 10 | | | | | | | | | | Male | 35-54 | White | |
| ER018 | Moderate | Sep-20 | Dec-20 | 95 | 117 | | | 12 | 19 | 83 | | | | | | | | | Female | 35-54 | White | |
| ER019 | Moderate | Sep-20 | --- | --- | | | | 12 | 19 | 83 | | | | | | | | | Male | 35-54 | White | |
| ER020 | Moderate | Sep-20 | Feb-21 | 154 | 175 | | | 82 | 188 | | | | | | | | | | Female | 55+ | White | |
| ER021 | Mild | Oct-20 | --- | --- | | | | 81 | | | | | | | | | | | Female | 18-34 | White | |
| ER022 | Mild | Nov-20 | --- | --- | | | | 7 | | | | | | | | | | | Female | 18-34 | American Indian or Alaska Native, Hispanic/Latino ethnicity | |
| ER023 | Moderate | Oct-20 | Jan-21 | 83 | 112 | | | 18 | 26 | 54 | 91 | | | | | | | | Female | 35-54 | Hispanic/Latino ethnicity | |
| ER024 | Mild | Nov-20 | Feb-21 | 84 | 120 | | | 79 | 182 | | | | | | | | | | Male | 35-54 | White and Hispanic/Latino ethnicity | |
| ER025 | Mild | Dec-20 | --- | --- | | | | 12 | 22 | 32 | 80 | | | | | | | | Male | 35-54 | White and Hispanic/Latino ethnicity | |
| ER026 | Mild | Jan-21 | --- | --- | | | | 27 | 48 | 76 | 138 | | | | | | | | Male | 35-54 | White/African American/American Indian/Alaskan race and Hispanic/Latino ethnicity | |
| ER027 | Mild | Nov-20 | Jan-21 | 73 | 96 | | | 5 | 26 | 75 | | | | | | | | | Female | 18-34 | Hispanic/Latino ethnicity | |
| ER028 | Moderate | Nov-20 | --- | --- | | | | 9 | 16 | 31 | 81 | | | | | | | | Male | 18-34 | American Indian/Aalaskan race and Hispanic/Latino ethnicity | |
| ER029 | Moderate | Nov-20 | --- | --- | | | | 2 | 10 | 33 | 75 | | | | | | | | Male | 55+ | White race/Hispanic/Latino ethnicity | |
| ER030 | Moderate | Nov-20 | Mar-21 | 124 | 152 | | | 10 | 16 | 32 | 82 | | | | | | | | Female | 18-34 | Hispanic/Latino ethnicity | |
| ER031 | Moderate | Nov-20 | May-21 | 168 | | | | 1 | 12 | 27 | 70 | | | | | | | | Female | 35-54 | white | |
| ER032 | Mild | Nov-20 | Dec-20 | 34 | 58 | | | 4 | 11 | 28 | 77 | | | | | | | | Male | 35-54 | white | |
| ER033 | MIld | Nov-20 | --- | --- | | | | 1 | 8 | | | | | | | | | | Male | 35-54 | . | |
| ER034 | Mild | Dec-20 | --- | --- | | | | 1 | | | | | | | | | | | Male | 18-34 | . | |
| ER035 | Moderate | Nov-20 | Feb-21 | 78 | 106 | | | 3 | 10 | 36 | 73 | | | | | | | | Female | 55+ | Hispanic/Latino ethnicity | |
| ER036 | Mild | Nov-20 | Jan-21 | 55 | 76 | | | 4 | 11 | 37 | 74 | | | | | | | | Female | 35-54 | Hispanic/Latino ethnicity | |
| ER037 | Mild | Dec-20 | Mar-21 | 93 | 123 | | | 4 | 12 | 34 | 70 | | | | | | | | Female | 18-34 | White race/Hispanic/Latino ethnicity | |
| ER038 | Moderate | Dec-20 | Jan-21 | 52 | 73 | | | 5 | 13 | 34 | 71 | | | | | | | | Male | 55+ | White race/Hispanic/Latino ethnicity | |
| ER039 | Moderate | Nov-20 | --- | --- | | | | 16 | 38 | 81 | | | | | | | | | Female | 55+ | . | |
| ER040 | Moderate | Dec-20 | Mar-21 | 84 | 112 | | | 2 | 15 | 31 | 74 | | | | | | | | Female | 55+ | Hispanic/Latino ethnicity | |
| ER041 | Moderate | Dec-20 | Mar-21 | 89 | 117 | | | 6 | 35 | 78 | | | | | | | | | Male | 55+ | Hispanic/Latino ethnicity | |
| ER042 | Moderate | Dec-20 | Jan-21 | 31 | 48 | | | 6 | 13 | 33 | 76 | | | | | | | | Female | 35-54 | Asian | |
| ER043 | Moderate | Dec-20 | Mar-21 | 84 | 112 | | | 3 | 14 | 30 | 73 | | | | | | | | Female | 35-54 | American indian/alaskan race and hispanic/latino ethnicity | |
| ER044 | Moderate | Dec-20 | Mar-21 | 79 | 108 | | | 4 | 17 | 24 | 75 | | | | | | | | Male | 55+ | White | |
| ER045 | Moderate | Jan-21 | Jan-21 | -2 | 43 | | | 3 | 12 | 26 | 75 | | | | | | | | Female | 35-54 | White and Hispanic/Latino ethnicity | |
| ER046 | Moderate | Jan-21 | Mar-21 | 69 | 97 | | | 3 | 12 | 26 | 75 | | | | | | | | Male | 35-54 | American Indian/Alaskan race and Hispanic/Latino ethnicity | |
| ER101 | --- | --- | Dec-20 | 0 (N/A) | 21 | 281 | 650 | | | | | | 92 | 306 | 634 | 663 | | | Female | 35-54 | Asian | |
| ER102 | --- | --- | May-21 | 0 (N/A) | 21 | | | | | | | | -75 | 14 | 36 | 56 | | | Male | 35-54 | African American | |
| ER103 | --- | --- | Mar-21 | 0 (N/A) | 19 | | | | | | | | 19 | | | | | | Female | 18-34 | Asian | |
| ER104 | --- | --- | --- | | | | | | | | | | --- | | | | | | Male | 18-34 | | |
| ER105 | --- | --- | Feb-21 | 0 (N/A) | 22 | 245 | 594 | | | | | | 377 | 611 | | | | | Female | 55+ | Asian | |
| ER106 | --- | --- | Jan-21 | 0 (N/A) | 28 | 282 | 597 | | | | | | 62 | 380 | 614 | | | | Male | 55+ | Asian | |
| ER107 | --- | --- | Feb-21 | 0 (N/A) | 21 | 324 | | | | | | | 0 | 19 | 42 | 64 | 323 | 417 | Female | 35-54 | White | |
| ER108 | --- | --- | Mar-21 | 0 (N/A) | 21 | 252 | 561 | | | | | | -3 | 20 | 250 | 277 | 553 | 581 | Male | 35-54 | Other | Additional sampling on 258, 422, 582 days post-vax |
| ER109 | --- | --- | Feb-21 | 0 (N/A) | 21 | 311 | | | | | | | -15 | 20 | 42 | 443 | 575 | 603 | Male | 18-34 | White | Additional sampling on 7, 589, 593, 604 days post-vax |
| ER110 | --- | --- | Mar-21 | 0 (N/A) | 21 | | | | | | | | -3 | 16 | 38 | | | | Male | 35-54 | White | |
| ER111 | --- | --- | --- | | | | | | | | | | --- | | | | | | Female | 35-54 | Hispanic/Latino ethnicity | |
| ER112 | --- | --- | --- | | | | | | | | | | --- | | | | | | Female | 35-54 | White | |
| ER113 | --- | --- | Mar-21 | 0 (N/A) | 21 | | | | | | | | 0 | 17 | 45 | | | | Male | 18-34 | White | |

### Slide 2
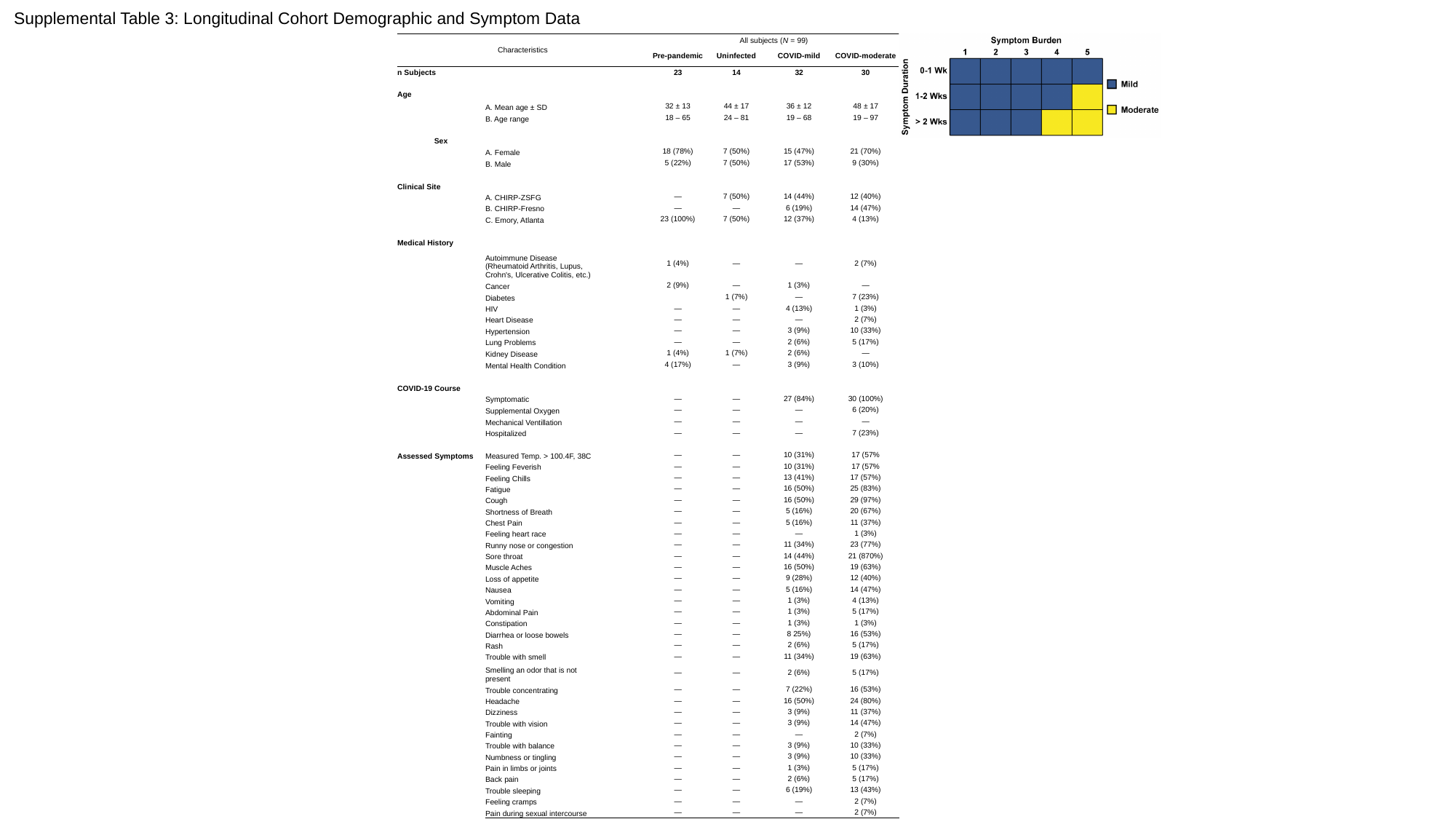

Supplemental Table 3: Longitudinal Cohort Demographic and Symptom Data
| Characteristics | | | All subjects (N = 99) | | | |
| --- | --- | --- | --- | --- | --- | --- |
| | | | Pre-pandemic | Uninfected | COVID-mild | COVID-moderate |
| n Subjects | | | 23 | 14 | 32 | 30 |
| Age | | | | | | |
| | A. Mean age ± SD | | 32 ± 13 | 44 ± 17 | 36 ± 12 | 48 ± 17 |
| | B. Age range | | 18 – 65 | 24 – 81 | 19 – 68 | 19 – 97 |
| Sex | | | | | | |
| | A. Female | | 18 (78%) | 7 (50%) | 15 (47%) | 21 (70%) |
| | B. Male | | 5 (22%) | 7 (50%) | 17 (53%) | 9 (30%) |
| Clinical Site | | | | | | |
| | A. CHIRP-ZSFG | | — | 7 (50%) | 14 (44%) | 12 (40%) |
| | B. CHIRP-Fresno | | — | — | 6 (19%) | 14 (47%) |
| | C. Emory, Atlanta | | 23 (100%) | 7 (50%) | 12 (37%) | 4 (13%) |
| Medical History | | | | | | |
| | Autoimmune Disease (Rheumatoid Arthritis, Lupus, Crohn's, Ulcerative Colitis, etc.) | | 1 (4%) | — | — | 2 (7%) |
| | Cancer | | 2 (9%) | — | 1 (3%) | — |
| | Diabetes | | | 1 (7%) | — | 7 (23%) |
| | HIV | | — | — | 4 (13%) | 1 (3%) |
| | Heart Disease | | — | — | — | 2 (7%) |
| | Hypertension | | — | — | 3 (9%) | 10 (33%) |
| | Lung Problems | | — | — | 2 (6%) | 5 (17%) |
| | Kidney Disease | | 1 (4%) | 1 (7%) | 2 (6%) | — |
| | Mental Health Condition | | 4 (17%) | — | 3 (9%) | 3 (10%) |
| COVID-19 Course | | | | | | |
| | Symptomatic | | — | — | 27 (84%) | 30 (100%) |
| | Supplemental Oxygen | | — | — | — | 6 (20%) |
| | Mechanical Ventillation | | — | — | — | — |
| | Hospitalized | | — | — | — | 7 (23%) |
| Assessed Symptoms | Measured Temp. > 100.4F, 38C | | — | — | 10 (31%) | 17 (57% |
| | Feeling Feverish | | — | — | 10 (31%) | 17 (57% |
| | Feeling Chills | | — | — | 13 (41%) | 17 (57%) |
| | Fatigue | | — | — | 16 (50%) | 25 (83%) |
| | Cough | | — | — | 16 (50%) | 29 (97%) |
| | Shortness of Breath | | — | — | 5 (16%) | 20 (67%) |
| | Chest Pain | | — | — | 5 (16%) | 11 (37%) |
| | Feeling heart race | | — | — | — | 1 (3%) |
| | Runny nose or congestion | | — | — | 11 (34%) | 23 (77%) |
| | Sore throat | | — | — | 14 (44%) | 21 (870%) |
| | Muscle Aches | | — | — | 16 (50%) | 19 (63%) |
| | Loss of appetite | | — | — | 9 (28%) | 12 (40%) |
| | Nausea | | — | — | 5 (16%) | 14 (47%) |
| | Vomiting | | — | — | 1 (3%) | 4 (13%) |
| | Abdominal Pain | | — | — | 1 (3%) | 5 (17%) |
| | Constipation | | — | — | 1 (3%) | 1 (3%) |
| | Diarrhea or loose bowels | | — | — | 8 25%) | 16 (53%) |
| | Rash | | — | — | 2 (6%) | 5 (17%) |
| | Trouble with smell | | — | — | 11 (34%) | 19 (63%) |
| | Smelling an odor that is not present | | — | — | 2 (6%) | 5 (17%) |
| | Trouble concentrating | | — | — | 7 (22%) | 16 (53%) |
| | Headache | | — | — | 16 (50%) | 24 (80%) |
| | Dizziness | | — | — | 3 (9%) | 11 (37%) |
| | Trouble with vision | | — | — | 3 (9%) | 14 (47%) |
| | Fainting | | — | — | — | 2 (7%) |
| | Trouble with balance | | — | — | 3 (9%) | 10 (33%) |
| | Numbness or tingling | | — | — | 3 (9%) | 10 (33%) |
| | Pain in limbs or joints | | — | — | 1 (3%) | 5 (17%) |
| | Back pain | | — | — | 2 (6%) | 5 (17%) |
| | Trouble sleeping | | — | — | 6 (19%) | 13 (43%) |
| | Feeling cramps | | — | — | — | 2 (7%) |
| | Pain during sexual intercourse | | — | — | — | 2 (7%) |

### Slide 3
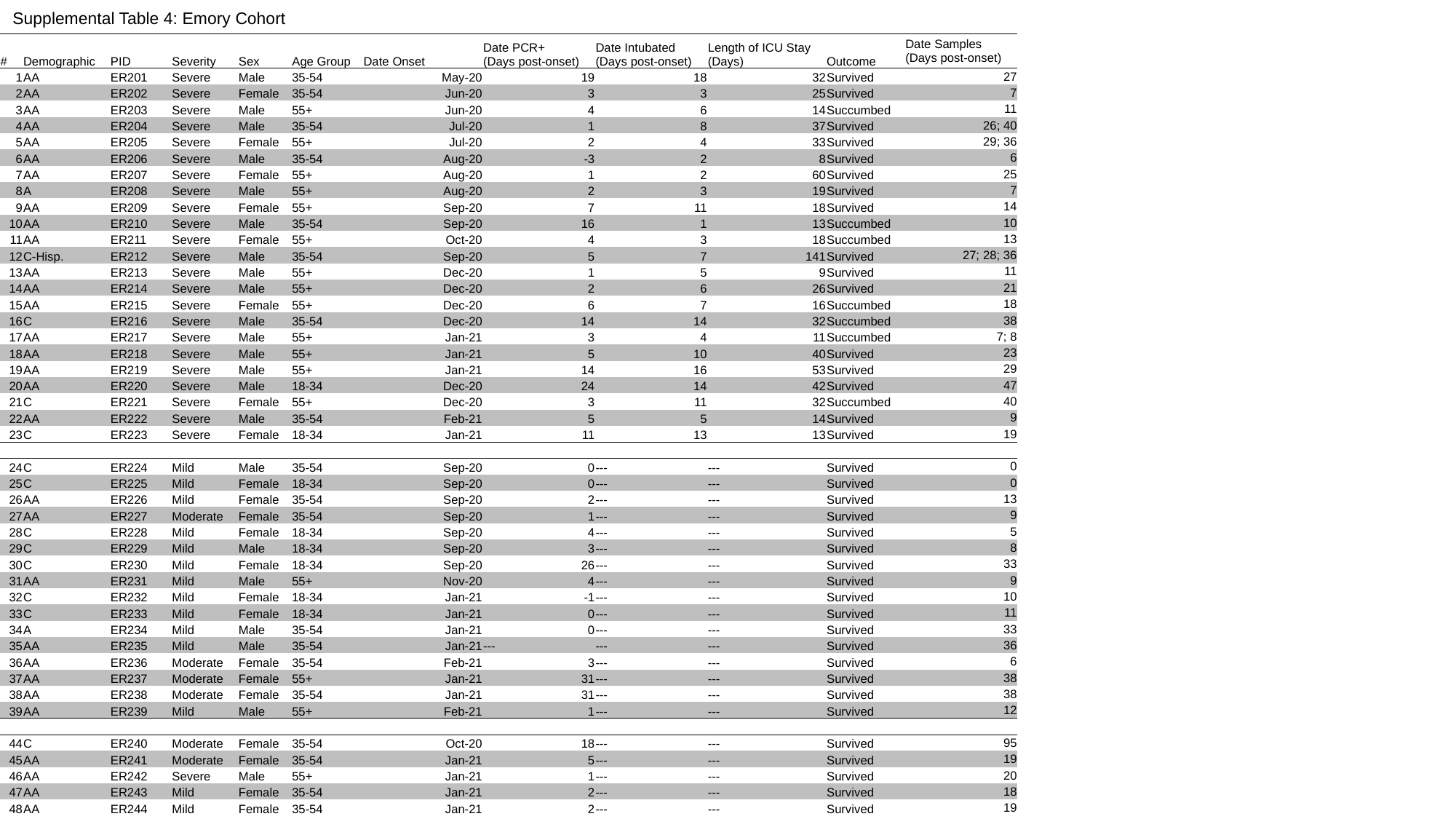

Supplemental Table 4: Emory Cohort
| # | Demographic | PID | Severity | Sex | Age Group | Date Onset | Date PCR+ (Days post-onset) | Date Intubated (Days post-onset) | Length of ICU Stay (Days) | Outcome | Date Samples (Days post-onset) |
| --- | --- | --- | --- | --- | --- | --- | --- | --- | --- | --- | --- |
| 1 | AA | ER201 | Severe | Male | 35-54 | May-20 | 19 | 18 | 32 | Survived | 27 |
| 2 | AA | ER202 | Severe | Female | 35-54 | Jun-20 | 3 | 3 | 25 | Survived | 7 |
| 3 | AA | ER203 | Severe | Male | 55+ | Jun-20 | 4 | 6 | 14 | Succumbed | 11 |
| 4 | AA | ER204 | Severe | Male | 35-54 | Jul-20 | 1 | 8 | 37 | Survived | 26; 40 |
| 5 | AA | ER205 | Severe | Female | 55+ | Jul-20 | 2 | 4 | 33 | Survived | 29; 36 |
| 6 | AA | ER206 | Severe | Male | 35-54 | Aug-20 | -3 | 2 | 8 | Survived | 6 |
| 7 | AA | ER207 | Severe | Female | 55+ | Aug-20 | 1 | 2 | 60 | Survived | 25 |
| 8 | A | ER208 | Severe | Male | 55+ | Aug-20 | 2 | 3 | 19 | Survived | 7 |
| 9 | AA | ER209 | Severe | Female | 55+ | Sep-20 | 7 | 11 | 18 | Survived | 14 |
| 10 | AA | ER210 | Severe | Male | 35-54 | Sep-20 | 16 | 1 | 13 | Succumbed | 10 |
| 11 | AA | ER211 | Severe | Female | 55+ | Oct-20 | 4 | 3 | 18 | Succumbed | 13 |
| 12 | C-Hisp. | ER212 | Severe | Male | 35-54 | Sep-20 | 5 | 7 | 141 | Survived | 27; 28; 36 |
| 13 | AA | ER213 | Severe | Male | 55+ | Dec-20 | 1 | 5 | 9 | Survived | 11 |
| 14 | AA | ER214 | Severe | Male | 55+ | Dec-20 | 2 | 6 | 26 | Survived | 21 |
| 15 | AA | ER215 | Severe | Female | 55+ | Dec-20 | 6 | 7 | 16 | Succumbed | 18 |
| 16 | C | ER216 | Severe | Male | 35-54 | Dec-20 | 14 | 14 | 32 | Succumbed | 38 |
| 17 | AA | ER217 | Severe | Male | 55+ | Jan-21 | 3 | 4 | 11 | Succumbed | 7; 8 |
| 18 | AA | ER218 | Severe | Male | 55+ | Jan-21 | 5 | 10 | 40 | Survived | 23 |
| 19 | AA | ER219 | Severe | Male | 55+ | Jan-21 | 14 | 16 | 53 | Survived | 29 |
| 20 | AA | ER220 | Severe | Male | 18-34 | Dec-20 | 24 | 14 | 42 | Survived | 47 |
| 21 | C | ER221 | Severe | Female | 55+ | Dec-20 | 3 | 11 | 32 | Succumbed | 40 |
| 22 | AA | ER222 | Severe | Male | 35-54 | Feb-21 | 5 | 5 | 14 | Survived | 9 |
| 23 | C | ER223 | Severe | Female | 18-34 | Jan-21 | 11 | 13 | 13 | Survived | 19 |
| 24 | C | ER224 | Mild | Male | 35-54 | Sep-20 | 0 | --- | --- | Survived | 0 |
| 25 | C | ER225 | Mild | Female | 18-34 | Sep-20 | 0 | --- | --- | Survived | 0 |
| 26 | AA | ER226 | Mild | Female | 35-54 | Sep-20 | 2 | --- | --- | Survived | 13 |
| 27 | AA | ER227 | Moderate | Female | 35-54 | Sep-20 | 1 | --- | --- | Survived | 9 |
| 28 | C | ER228 | Mild | Female | 18-34 | Sep-20 | 4 | --- | --- | Survived | 5 |
| 29 | C | ER229 | Mild | Male | 18-34 | Sep-20 | 3 | --- | --- | Survived | 8 |
| 30 | C | ER230 | Mild | Female | 18-34 | Sep-20 | 26 | --- | --- | Survived | 33 |
| 31 | AA | ER231 | Mild | Male | 55+ | Nov-20 | 4 | --- | --- | Survived | 9 |
| 32 | C | ER232 | Mild | Female | 18-34 | Jan-21 | -1 | --- | --- | Survived | 10 |
| 33 | C | ER233 | Mild | Female | 18-34 | Jan-21 | 0 | --- | --- | Survived | 11 |
| 34 | A | ER234 | Mild | Male | 35-54 | Jan-21 | 0 | --- | --- | Survived | 33 |
| 35 | AA | ER235 | Mild | Male | 35-54 | Jan-21 | --- | --- | --- | Survived | 36 |
| 36 | AA | ER236 | Moderate | Female | 35-54 | Feb-21 | 3 | --- | --- | Survived | 6 |
| 37 | AA | ER237 | Moderate | Female | 55+ | Jan-21 | 31 | --- | --- | Survived | 38 |
| 38 | AA | ER238 | Moderate | Female | 35-54 | Jan-21 | 31 | --- | --- | Survived | 38 |
| 39 | AA | ER239 | Mild | Male | 55+ | Feb-21 | 1 | --- | --- | Survived | 12 |
| 44 | C | ER240 | Moderate | Female | 35-54 | Oct-20 | 18 | --- | --- | Survived | 95 |
| 45 | AA | ER241 | Moderate | Female | 35-54 | Jan-21 | 5 | --- | --- | Survived | 19 |
| 46 | AA | ER242 | Severe | Male | 55+ | Jan-21 | 1 | --- | --- | Survived | 20 |
| 47 | AA | ER243 | Mild | Female | 35-54 | Jan-21 | 2 | --- | --- | Survived | 18 |
| 48 | AA | ER244 | Mild | Female | 35-54 | Jan-21 | 2 | --- | --- | Survived | 19 |

### Slide 4
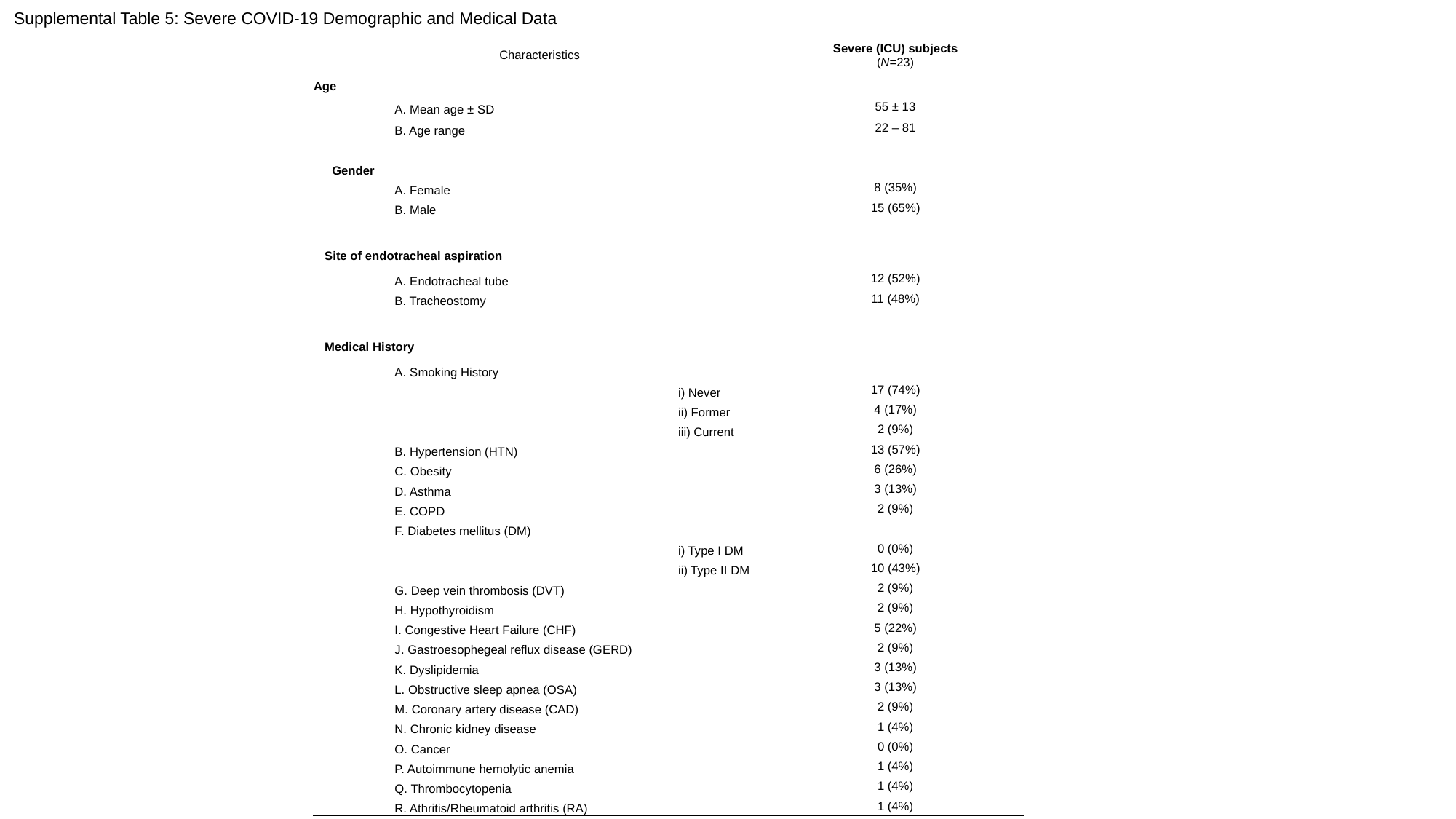

Supplemental Table 5: Severe COVID-19 Demographic and Medical Data
| Characteristics | | | Severe (ICU) subjects (N=23) |
| --- | --- | --- | --- |
| Age | | | |
| | A. Mean age ± SD | | 55 ± 13 |
| | B. Age range | | 22 – 81 |
| Gender | | | |
| | A. Female | | 8 (35%) |
| | B. Male | | 15 (65%) |
| Site of endotracheal aspiration | | | |
| | A. Endotracheal tube | | 12 (52%) |
| | B. Tracheostomy | | 11 (48%) |
| Medical History | | | |
| | A. Smoking History | | |
| | | i) Never | 17 (74%) |
| | | ii) Former | 4 (17%) |
| | | iii) Current | 2 (9%) |
| | B. Hypertension (HTN) | | 13 (57%) |
| | C. Obesity | | 6 (26%) |
| | D. Asthma | | 3 (13%) |
| | E. COPD | | 2 (9%) |
| | F. Diabetes mellitus (DM) | | |
| | | i) Type I DM | 0 (0%) |
| | | ii) Type II DM | 10 (43%) |
| | G. Deep vein thrombosis (DVT) | | 2 (9%) |
| | H. Hypothyroidism | | 2 (9%) |
| | I. Congestive Heart Failure (CHF) | | 5 (22%) |
| | J. Gastroesophegeal reflux disease (GERD) | | 2 (9%) |
| | K. Dyslipidemia | | 3 (13%) |
| | L. Obstructive sleep apnea (OSA) | | 3 (13%) |
| | M. Coronary artery disease (CAD) | | 2 (9%) |
| | N. Chronic kidney disease | | 1 (4%) |
| | O. Cancer | | 0 (0%) |
| | P. Autoimmune hemolytic anemia | | 1 (4%) |
| | Q. Thrombocytopenia | | 1 (4%) |
| | R. Athritis/Rheumatoid arthritis (RA) | | 1 (4%) |
