## Supplemental Figures S1-S6 and Table S1 for "Transient anti-interferon autoantibodies in the airways are associated with efficient recovery from COVID-19"

| Ag # | Ag Class | Protein | MW (kDa) | Source | Cell Line | Cat# |
|---|---|---|---|---|---|---|
| 1 | SCV2 Structural | RBD | 30 | GenScript | HEK293 | Z03483-1 |
| 2 | SCV2 Structural | Spike S1 | 76.9 | Acro Biosystems | HEK293 | S1N-C52H3 |
| 3 | SCV2 Structural | Spike S2 | 60 | Acro Biosystems | HEK293 | S2N-C52H5 |
| 4 | SCV2 Structural | NP (ORF9a) | 47 | RayBioTech | HEK293 | 230-30164-100 |
| 5 | SCV2 Structural | E (ORF4) | 50.2 | Chempartner | HEK293 | |
| 6 | SCV2 Structural | M (ORF5) | 66.9 | Chempartner | HEK293 | |
| 7 | SCV2 Nonstructural | ORF3a | 34.4 | Chempartner | HEK293 | |
| 8 | SCV2 Nonstructural | ORF3b | 48.3 | Chempartner | HEK293 | |
| 9 | SCV2 Nonstructural | ORF8 | 15.4 | Chempartner | HEK293 | |
| 10 | SCV2 Nonstructural | ORF9b | 14.2 | Chempartner | HEK293 | |
| 11 | SCV2 Nonstructural | NSP1 | 23.2 | Chempartner | HEK293 | |
| 12 | SCV2 Nonstructural | NSP2 | 74 | Chempartner | HEK293 | |
| 13 | SCV2 Nonstructural | NSP7 | 12.812 | Chempartner | HEK293 | |
| 14 | SCV2 Nonstructural | NSP8 | 25.453 | Chempartner | HEK293 | |
| 15 | SCV2 Nonstructural | NSP12 | 110.03 | Chempartner | HEK293 | |
| 16 | Autoreactive | IFNomega | 19.9 | Sigma Aldrich | E. Coli | SRP3061 |
| 17 | Autoreactive | IFNa2a | 19.2 | Miltenyi | E. Coli | 130-093-874 |
| 18 | Autoreactive | 9G4 | 150 | Sanz Lab | N/A | |
| 19 | Internal Control | BSA | 66 | Sigma-Aldrich | N/A | B6917 |
| 20 | VOC Structural | BA.4/5 NP | 47.1 | Acro Biosystems | HEK293 | NUN-C52Hx |
| 21 | VOC Structural | BA.4/5 RBD | 28.3 | Acro Biosystems | HEK293 | SPD-C82Ew |
| 22 | VOC Structural | Delta RBD | 26.6 | Sino Biological | HEK293 | 40592-V08H90 |

**Table S1: FlowBEAT antigens**

List of all 22 flowBEAT antigens, including SARS-CoV-2 structural and non-structural viral antigens, internal controls, human autoantigens and SARS-CoV-2 variant antigens.

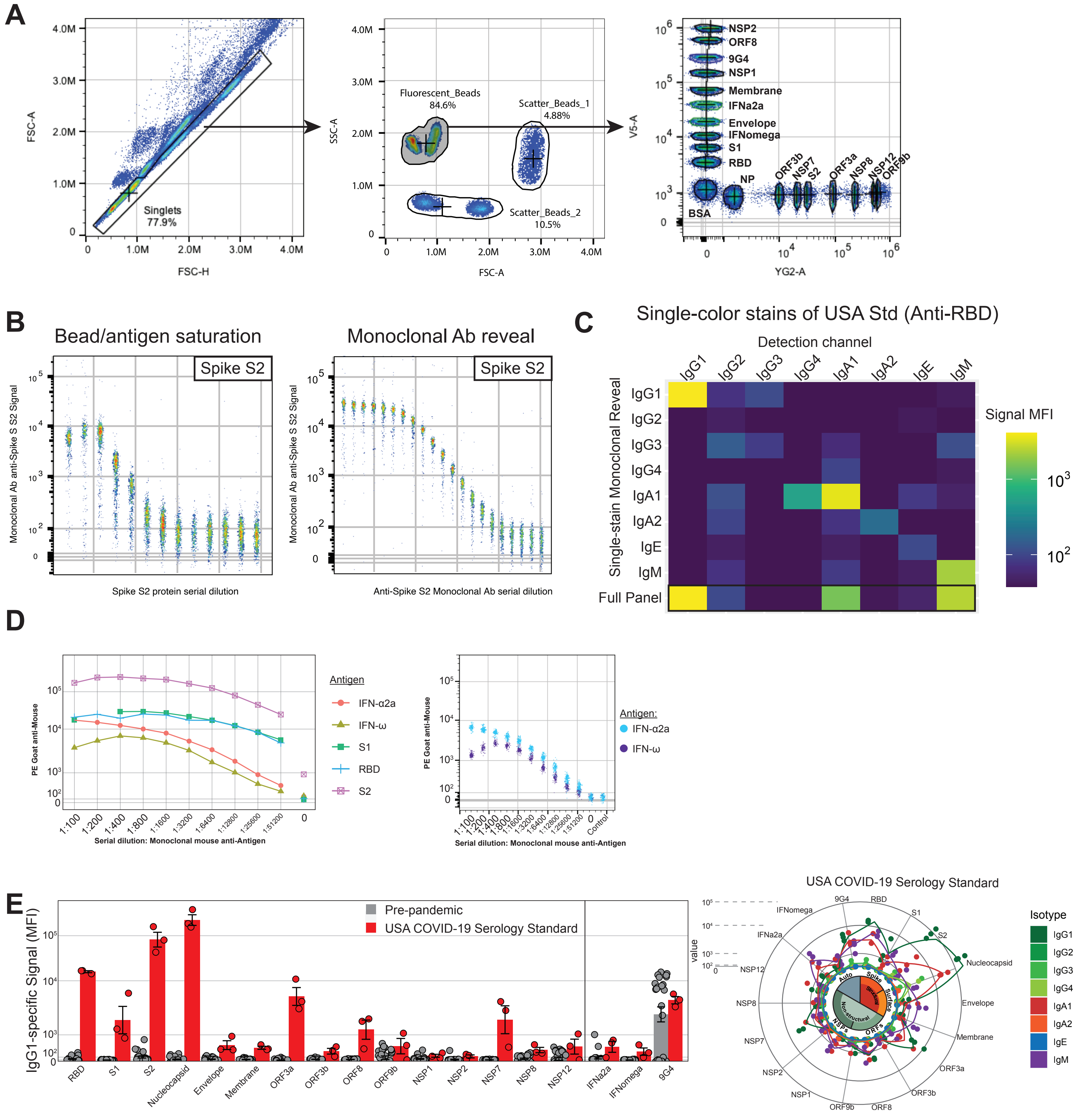
A
FSC-A
FSC-H
Singlets
77.9%
SSC-A
FSC-A
Fluorescent_Beads
84.6%
Scatter_Beads_1
4.88%
Scatter_Beads_2
10.5%
V5-A
YG2-A
NSP2
ORF8
9G4
NSP1
Membrane
IFNa2a
Envelope
IFNomega
S1
RBD
NP
BSA
ORF3b
NSP7
S2
ORF3a
NSP8
NSP12
ORF9b
B
Bead/antigen saturation
Monoclonal Ab reveal
Spike S2
Monoclonal Ab anti-Spike S S2 Signal
Spike S2 protein serial dilution
Anti-Spike S2 Monoclonal Ab serial dilution
C
Single-color stains of USA Std (Anti-RBD)
Detection channel
IgG1 IgG2 IgG3 IgG4 IgA1 IgA2 IgE IgM
Single-stain Monoclonal Reveal
Full Panel
Signal MFI
D
PE Goat anti-Mouse
Antigen
IFN-α2a
IFN-ω
S1
RBD
S2
Serial dilution: Monoclonal mouse anti-Antigen
Antigen:
Control
E
IgG1-specific Signal (MFI)
Pre-pandemic
USA COVID-19 Serology Standard
RBD S1 S2 Nucleocapsid Envelope Membrane ORF3a ORF3b ORF8 ORF9b NSP1 NSP2 NSP7 NSP8 NSP12 IFNa2a IFNomega 9G4
value
Auto
Spike
Surface
Structural
Non-structural
NSPs
ORFs
Isotype
IgG1 IgG2 IgG3 IgG4 IgA1 IgA2 IgE IgM

**Fig. S1: FlowBEAT validation and reproducibility**

(**A**) FlowBEAT gating strategy for 19 fluorescent + 3 scatter-discriminated bead species (22 bead species). (**B**) Leftmost plot: determination of bead-antigen saturation point by serial dilution (X-axis) of biotin-conjugated spike S2 antigen in the presence of a constant amount of beads in constant volume. Reveal signal generated by monoclonal mouse anti-S2 antibody with a secondary goat anti-mouse fluorescence. Rightmost plot: determination of sensitivity by serial dilution of monoclonal mouse anti-S2 antibody, serially diluted (X-axis) and staining S2-saturated beads, revealed by a secondary goat anti-mouse fluorescence. (**C**) Single-color stains reveal isotype specificity of flowBEAT assay. (**D**) Antigen-specificity demonstrated by antigen-specific monoclonal mouse anti-IFN-ω, anti-IFN-α2a, and anti-spike RBD, S1, S2 revealed by PE goat anti-mouse (Y-axis). (**E**) Bar plot of IgG1 anti-SARS-CoV-2 and autoantigens between pre-pandemic (grey) and USA SARS-CoV-2 serology standard (red). (Sunburst plot demonstrates total antigen reactivity in three independent replicates of USA SARS-CoV-2 serology standard.

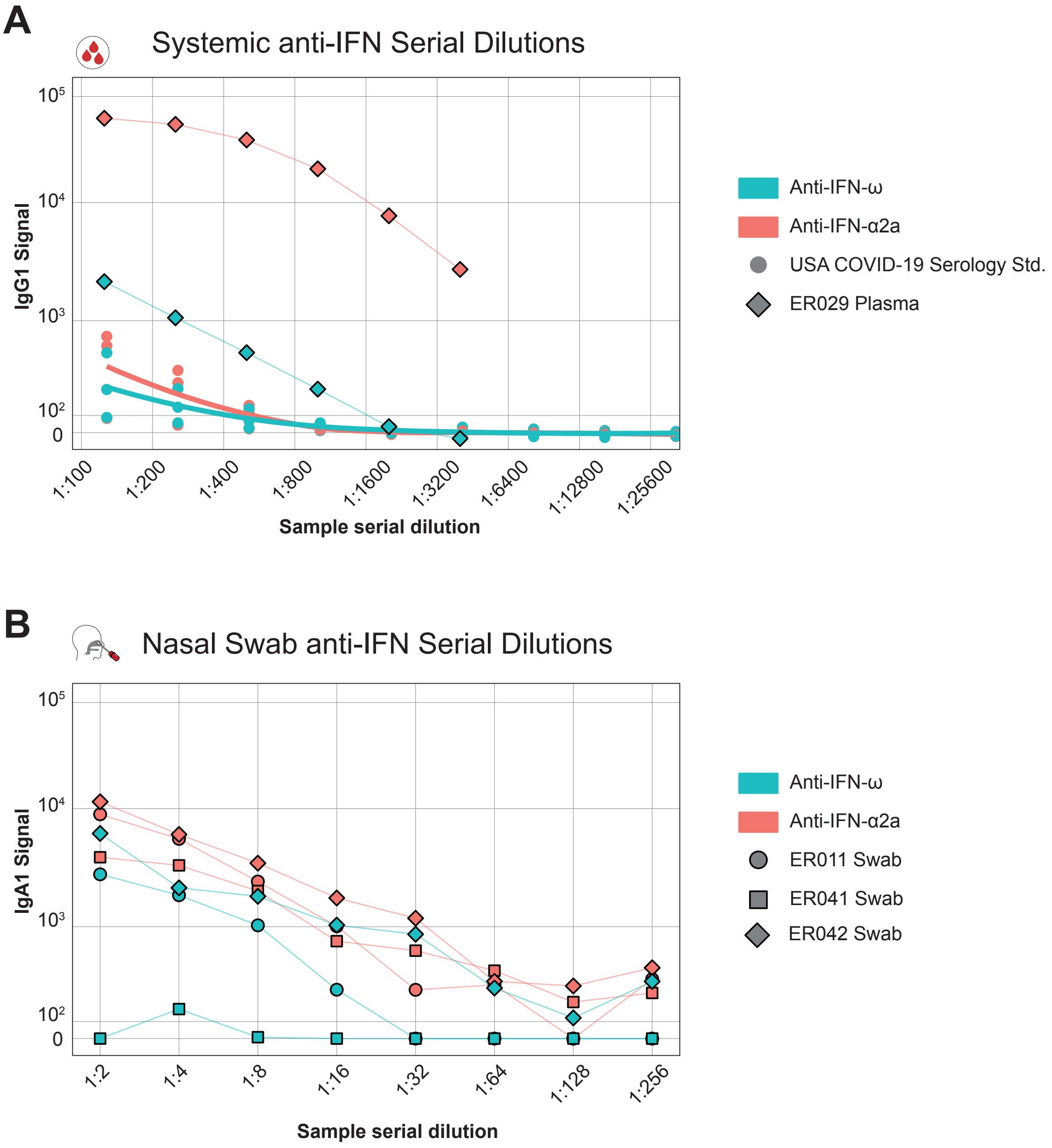
A
Systemic anti-IFN Serial Dilutions
IgG1 Signal
10^5
10^4
10^3
10^2
0
1:100
1:200
1:400
1:800
1:1600
1:3200
1:6400
1:12800
1:25600
Sample serial dilution
Anti-IFN-ω
Anti-IFN-α2a
USA COVID-19 Serology Std.
ER029 Plasma
B
Nasal Swab anti-IFN Serial Dilutions
IgA1 Signal
10^5
10^4
10^3
10^2
0
1:2
1:4
1:8
1:16
1:32
1:64
1:128
1:256
Sample serial dilution
Anti-IFN-ω
Anti-IFN-α2a
ER011 Swab
ER041 Swab
ER042 Swab

**Fig. S2: Serial dilution of anti-IFN-α2a positive samples to validate anti-IFN-α2a and anti-IFN-ω signals**
(**A**) Serial dilution of mild-infected blood anti-IFN IgG1 signals from one donor, and triplicate dilutions of blood anti-IFN signals from the USA serology standard. (**B**) Serial dilution of mild-infected nasal anti-IFN IgA1 signals.

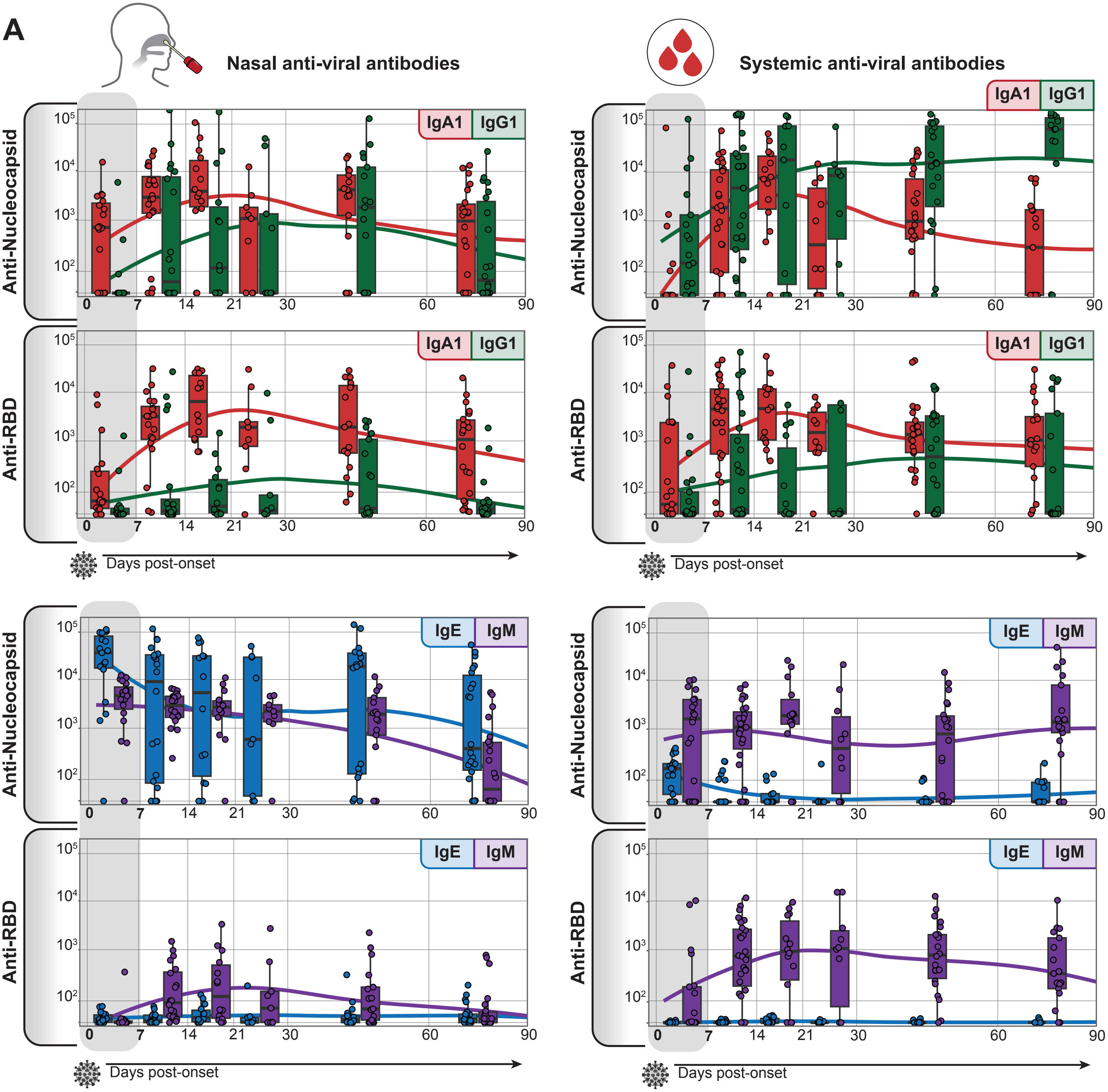
A
Nasal anti-viral antibodies
Systemic anti-viral antibodies
Anti-Nucleocapsid
Anti-RBD
IgA1
IgG1
IgE
IgM
Days post-onset
10⁵
10⁴
10³
10²
0
7
14
21
30
60
90

**Fig. S3: Antibody isotype response to COVID-19 infection +/-  vaccination in airway mucosa and blood**
(**A**) Box plots (binned by weeks post-onset) and trendlines (drawn from non-binned data) for mild- and moderate-infected airway and systemic anti-RBD and anti-Nucleocapsid signals, with IgA1 and IgG1 or IgE and IgM values plotted, as indicated.

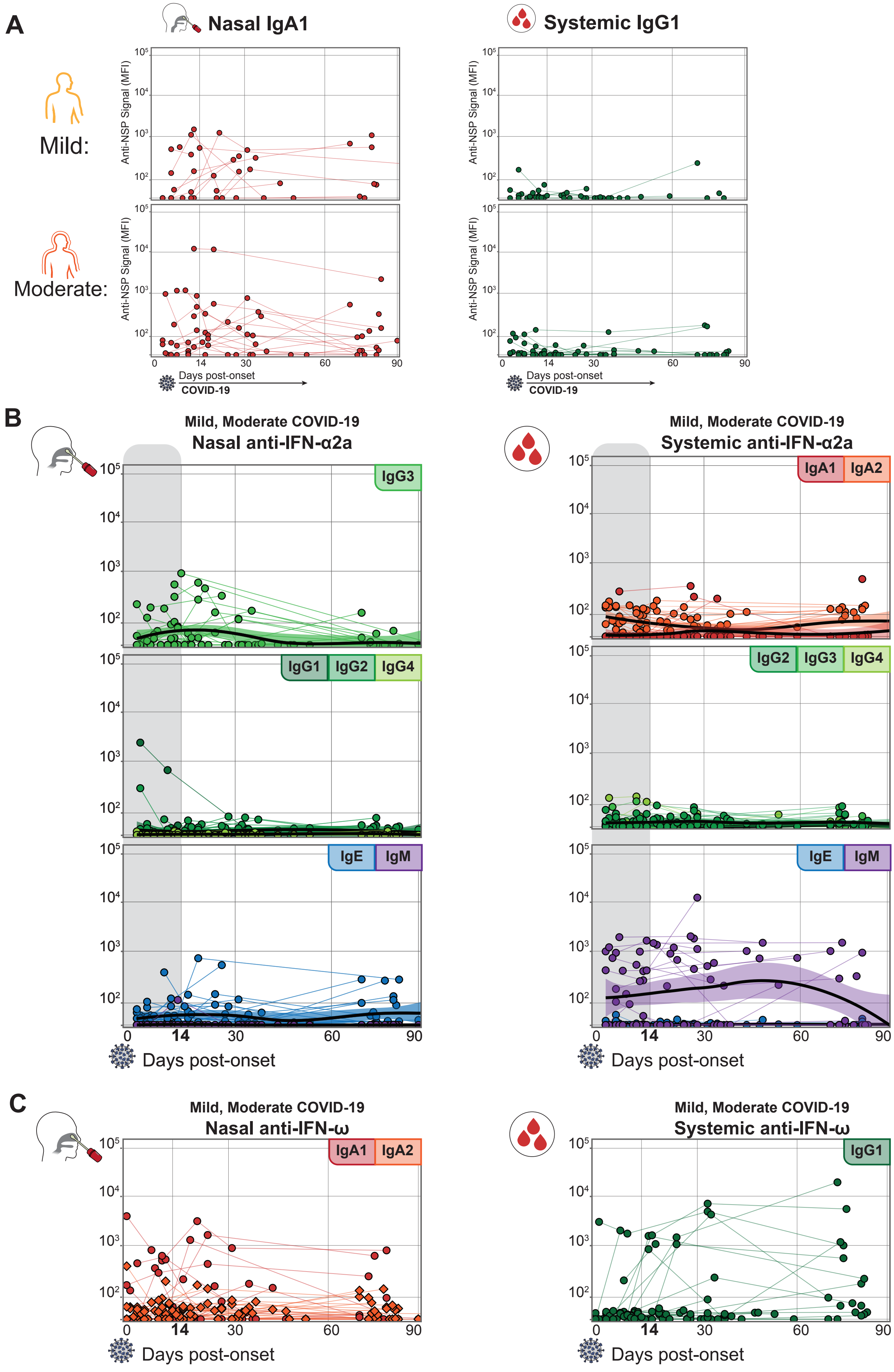

A
Nasal IgA1
Systemic IgG1
Mild:
Moderate:
Anti-NSP Signal (MFI)
Days post-onset
COVID-19
B
Mild, Moderate COVID-19
Nasal anti-IFN-α2a
IgG3
IgG1
IgG2
IgG4
IgE
IgM
Days post-onset
Mild, Moderate COVID-19
Systemic anti-IFN-α2a
IgA1
IgA2
IgG2
IgG3
IgG4
IgE
IgM
Days post-onset
C
Mild, Moderate COVID-19
Nasal anti-IFN-ω
IgA1
IgA2
Days post-onset
Mild, Moderate COVID-19
Systemic anti-IFN-ω
IgG1
Days post-onset

**Fig. S4: Anti-NSP nasal-specific programs and additional induced isotypes of anti-IFN-α2a**

(**A**) Anti-NSP7, NSP8 and ORF9a signals comprise a nasal-specific anti-NSP antibody response rarely detected in the blood of mild and moderate COVID-19 donors. (**B**) Additional longitudinal plots of anti-IFN-α2a-specific isotypes of nasal and systemic autoantibodies depict isotypes not displayed in Fig. 3A and 3C. Longitudinal samples of individual donors are linked by solid lines. Loess regressions plotted as solid black lines with shaded standard error. (**C**) Longitudinal plots of anti-IFN-ω-specific isotypes of nasal and systemic autoantibodies.

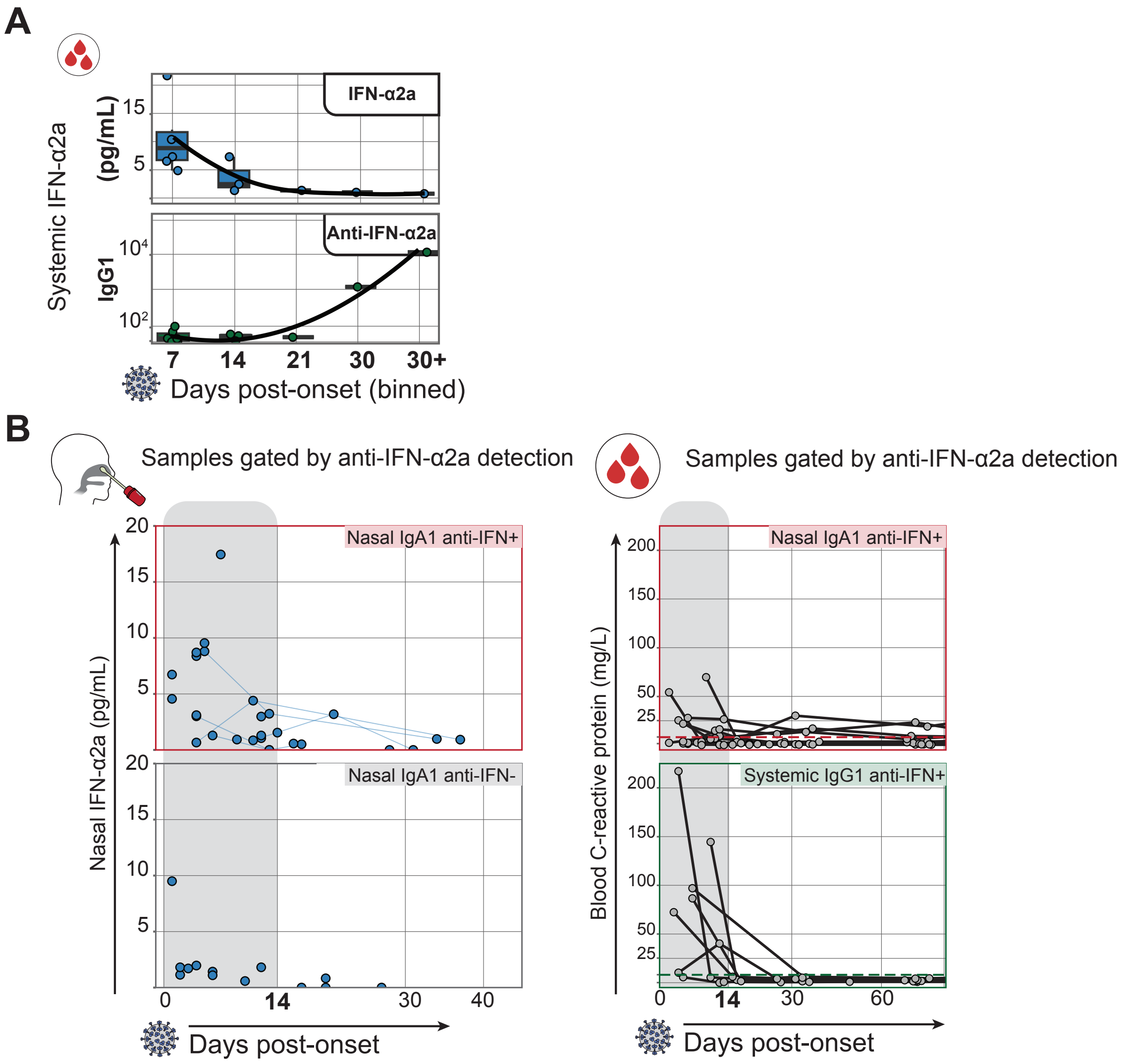

A
IFN-α2a
Anti-IFN-α2a
Systemic IFN-α2a
(pg/mL)
IgG1
15
5
$10^4$
$10^2$
7
14
21
30
30+
Days post-onset (binned)
B
Samples gated by anti-IFN-α2a detection
Samples gated by anti-IFN-α2a detection
Nasal IgA1 anti-IFN+
Nasal IgA1 anti-IFN-
Nasal IFN-α2a (pg/mL)
20
15
10
5
0
14
30
40
Days post-onset
Nasal IgA1 anti-IFN+
Systemic IgG1 anti-IFN+
Blood C-reactive protein (mg/L)
200
150
100
50
25
0
14
30
60
Days post-onset

**Fig. S5: Signals of inflammation and viral load in mild and moderate COVID-19 anti-IFN-α2a producers**
(**A**) Systemic IFN-α2a cytokine levels and corresponding systemic anti-IFN-α2a from the same samples. Points binned by days post-onset, as indicated in X-axis. (**B**) Longitudinal plots of nasal IFN-α2a cytokine (nasal, left) and systemic CRP (systemic, right), divided by nasal IgA1 anti-IFN-α2a-positive (left, red) and nasal IgA1 anti-IFN-α2a-negative (left, gray) or nasal IgA1 anti-IFN-α2a-positive (right, red) and systemic IgG1 anti-IFN-α2a-positive (right, green). Gates for relevant populations (red, green, gray color-coded) depicted in Fig. 3C.

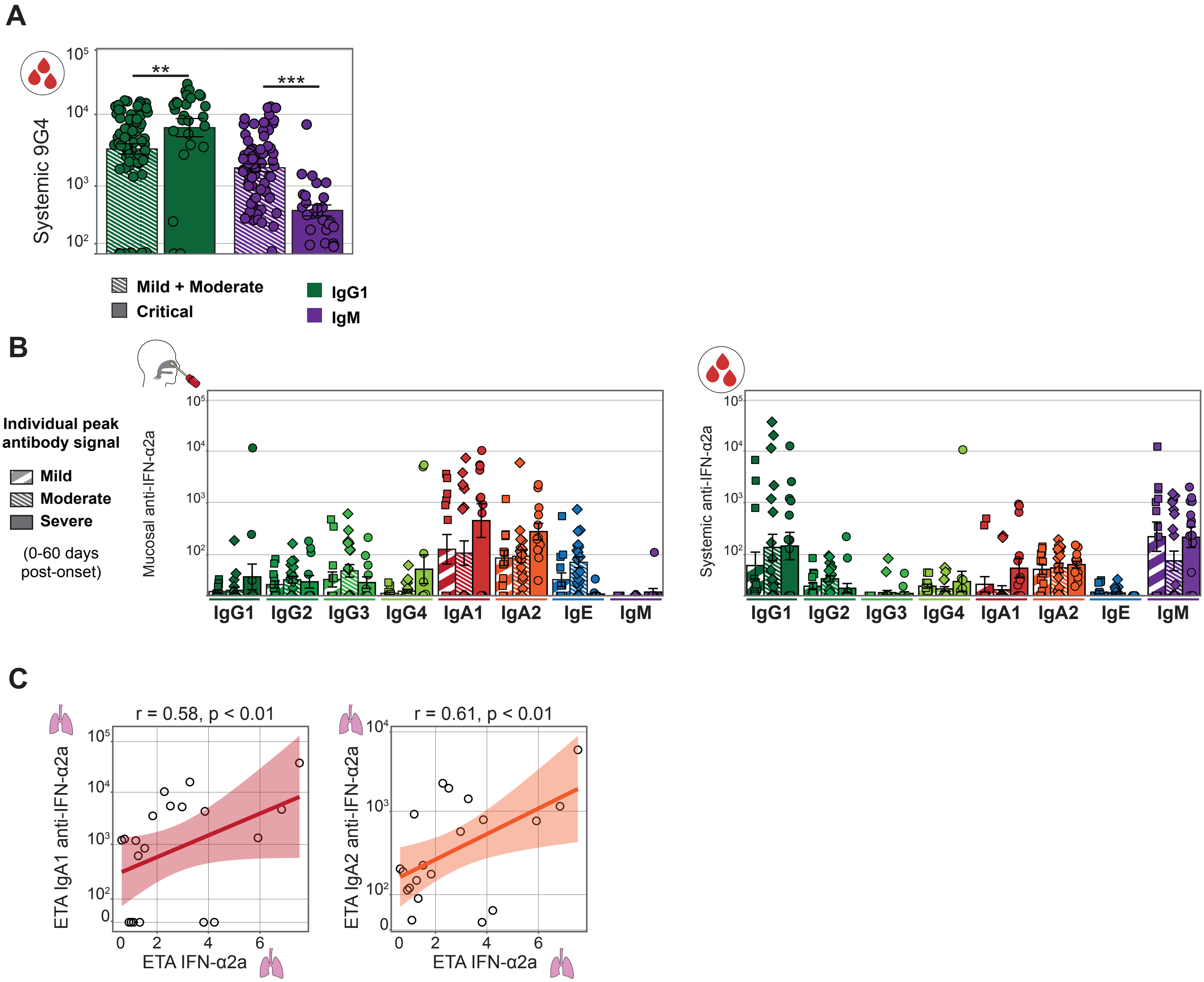

A
Systemic 9G4
**
***
Mild + Moderate
Critical
IgG1
IgM
B
Individual peak antibody signal
Mild
Moderate
Severe
(0-60 days post-onset)
Mucosal anti-IFN-α2a
Systemic anti-IFN-α2a
IgG1
IgG2
IgG3
IgG4
IgA1
IgA2
IgE
IgM
C
r = 0.58, p < 0.01
r = 0.61, p < 0.01
ETA IgA1 anti-IFN-α2a
ETA IgA2 anti-IFN-α2a
ETA IFN-α2a

**Fig. S6: Additional isotypes of autoreactive antibody response in mild, moderate, and severe COVID-19**
(**A**) Bar plot of systemic VH4-34 antibody detection (Y-axis) divided by isotype (color) and COVID-19 severity (pattern). Signal depicted is maximum signal detected over acute COVID-19 infection. Wilcoxon rank-sum test, $p < 0.01$, $p < 0.001$ as indicated. (**B**) Bar plot of systemic and airway anti-IFN-α2a (Y-axis) divided by isotype (color) and COVID-19 severity (pattern). Signal depicted is maximum signal detected over acute COVID-19 infection. (**C**) Correlation plots of ETA anti-IFN-α2a and ETA IFN-α2a cytokine levels in critical COVID-19 patients. Pearson product moment correlation coefficient, $p < 0.01$.
